# Automated quantification of Ki-67 expression in breast cancer from H&E-stained slides using a transformer-based regression model

**DOI:** 10.1101/2025.04.28.25326603

**Authors:** Abadh K Chaurasia, Patrick W Toohey, Matthew T Bennett, Helen C Harris, Alex W Hewitt

## Abstract

**Background:** Accurate quantification of the Ki-67 proliferation index is essential for breast cancer prognosis and treatment planning. Current automated methods, including classical and deep learning approaches based on cell detection or segmentation, often face challenges due to densely packed nuclei, morphological variability, and inter-laboratory differences. Since Hematoxylin and Eosin (H&E) staining is routinely performed, accurately estimating Ki-67 from these slides could save resources by eliminating the need for additional immunohistochemical (IHC) staining. We developed and validated a transformer-based regression model to estimate Ki-67 expression directly from H&E-stained Whole Slide Images (WSIs).

**Methods:** We used seven public datasets to select optimal transformer-based architectures and hyperparameters. WSIs underwent preprocessing to filter poor-quality patches, with a classification model identifying gradable patches. Only gradable patches proceeded to Ki-67 quantification. Initially, a regression model was trained on IHC-stained patches using independently annotated datasets, bypassing segmentation methods. This model generated pseudo-labels for unlabeled IHC patches, which were then paired with corresponding H&E images, with a separate model trained using only these H&E patches. Both models were evaluated separately across 1153 H&E and 843 IHC-stained WSIs, employing metrics such as R².

**Results:** Our regression model had good predictive accuracy, with R² values exceeding 0.90 for quantifying positive cells, negative cells, and Ki-67 ratios. The classification model effectively distinguished gradable patches, achieving a near-perfect AUROC (∼100%) across independent and unseen datasets. Cross-modality performance was robust, achieving R² values over 0.95 for positive and negative cell counts. Additionally, the model accurately captured the proliferation patterns from H&E-stained WSIs.

**Conclusion:** Our approach precisely quantifies Ki-67 expression and automates hotspot detection from WSIs, providing a scalable tool for digital pathology workflows. The cross-modality model can quantify molecular expression from morphological features using H&E-stained patches.

## INTRODUCTION

Breast cancer is a primary health concern worldwide, being the second most diagnosed cancer and the leading cause of cancer-related mortality among women.^1,2^ It is estimated to increase over three million new cases and one million deaths in 2040, with incidence rates varying widely by geographic region.^2^ The prevalence of breast cancer is highest in high-income regions but with lower mortality due to early detection and better healthcare, while transitioning countries, particularly in Africa and Asia, are experiencing rising incidence and disproportionately high mortality due to limited access to healthcare, late diagnoses, and inadequate treatment facilities.^1,3^ The factors contributing to breast cancer risk are complex and multifaceted, involving lifestyle choices, genetic predispositions, and environmental influences.^4,5^ Addressing global disparities in breast cancer and its complexities requires a comprehensive strategy that raises cancer awareness, improves healthcare access, and strengthens health systems with cutting-edge technology to support early detection and effective management of breast cancer globally.

The nuclear protein Ki-67 is an important prognostic and predictive biomarker for determining cellular proliferation in cancer, with high levels indicating a more aggressive disease and a poorer prognosis.^6,7^ The gold standard for quantifying the Ki-67 index involves manual pathologists assessing at least 500 malignant cells through immunohistochemistry (IHC), estimating the percentage of tumour cells expressing the Ki-67 antigen.^8–10^ Typically, pathologists visually analyse IHC-stained tumour specimens under a microscope to determine the percentage of Ki-67-positive cells, but this inherently subjective process can lead to significant variability in scores between different observers.^11^ This variability undermines the reliability of the Ki-67 index as a prognostic marker, impacting clinical decisions regarding treatment intensity and patient management. The lack of standardisation in Ki-67 evaluation has long been a concern in histopathology,^9,10^ highlighting the necessity for more objective, reproducible, and automated methods to ensure accurate estimates of the Ki-67 index, which currently limits its clinical utility in breast cancer.

Recent advancements in digital pathology and artificial intelligence (AI) have improved the extraction of tissue morphometric features, enhancing biomarker development in clinical oncology.^12–14^ Digital pathology provides high-resolution images, while deep learning models, including convolutional neural networks (CNN) and vision transformers (ViT), outperform in automating histopathological tasks, including nuclei segmentation and tumour grading.^12,15–17^ While multiple automated approaches have been applied, from classical machine learning to deep learning models based on cell detection or segmentation, these methods often struggle with densely packed or inadequately stained nuclei, morphological variability, and inter-laboratory heterogeneity.^13,18–20^ Thus, to overcome these challenges, we must select the optimal architectures, hyperparameters, and multi-centred data to develop a robust model that learns the inherent features from the nuclei, and it automatically determines the cut-off threshold for distinguishing between positive and negative-stained cells without relying on segmentation techniques.

ViT-based models outperform traditional CNN-based models in medical image analysis by providing superior contextual understanding, robustness, computational efficiency, and accuracy.^21^ Our study presents a novel, dual-stage AI-based framework for quantifying the Ki-67 index from Hematoxylin and Eosin (H&E)-stained Whole-Slide Images (WSIs). It utilises a ViT-based regression model for automatic nucleus detection and stain-invariant feature extraction from diverse data, without employing traditional approaches. In the first stage, we trained a regression model on IHC patches (manually graded by pathologists) to accurately quantify Ki-67 expression. We then applied a cross-stained pseudo-labelling strategy to quantify the molecular expression from morphological features using H&E-stained patches. Additionally, we develop a classification model to identify gradable histological patches from WSIs, ensuring that the regression models accurately estimate the Ki-67 index from clinically relevant tissue patches. This approach precisely estimates the Ki-67 index from WSIs and allowing for direct integration into digital pathology workflows.

## METHODS

### Study Overview

This study offers an automated, AI-driven system for quantifying the Ki-67 proliferation index and visualising hotspots (high densities of Ki-67-positive cells) from histological slides in breast cancer. The workflow comprises WSIs preprocessing, patch extraction, filtering of extracted gradable patches, and performing regression analysis to quantify the Ki-67 index from the WSIs.

All statistical analyses and training procedures were conducted in a virtual cloud environment using an NVIDIA GPU (40 GB RAM) with the Fastai (2.7.18) framework, the PyTorch (2.5.1+cu124) library, and the Python programming language (3.10.12).^22–24^

### Ethics Statement and Datasets

We incorporated seven publicly accessible histopathology datasets without direct patient interaction or personally identifiable information; no additional ethical approval was required. It complies with ethical guidelines, and all dataset sources follow data-sharing policies. A complete description of the contributing datasets is contained in the **Supplementary Methods**, and **Table 1** displays the composition of patches or WSIs used in this study.

**Table 1.** Datasets used in this study for the model development, validation, and external testing.

| **Dataset Name** | **Patches** | **Positive cells** | **Negative cells** |
| --- | --- | --- | --- |
| SHIDC-B-Ki-67<sup>25</sup> | 2356 | 50861 | 107653 |
| BCData<sup>26</sup> | 1338 | 62623 | 118451 |
| DeepSlides<sup>27</sup> | 694 | - | - |
| IHC4BC<sup>28</sup> | 20656 | - | - |
| ACROBAT<sup>29</sup> | 1996* | - | - |
| TCGA-UT<sup>30</sup> | 3561** | - | - |
| SICAP<sup>31</sup> | 133** | - | - |

\* 1153 H&E and 843 IHC-stained Whole Slide Images. \*\*Random patches were selected from the total dataset.

### Data Preparation for Regression Model

Developing a regression model for assessing the Ki-67 proliferation index from IHC-stained patches, we utilised two publicly available labelled datasets for Ki-67 expression: SHIDC-B-Ki-67 and BCData. Both datasets included high-resolution histopathology patches with annotated positive and negative cell counts. In the SHIDC-B-Ki-67 dataset, each image was paired with a corresponding file containing annotations for individual cells.^32^ We parsed each file to identify and count the number of positive and negative cells per image. The Ki-67 index was then computed based on the number of positive and negative cells in each patch.

The BCData dataset includes patches with annotated coordinates for positive and negative cells.^26^ We extracted these annotations using the h5py library^33^, counted the positive and negative cells for each patch, and then calculated the Ki-67 index. We compiled the data into a structured tabular format for both datasets, including the image path and positive and negative cell counts.

In addition to traditional augmentation during training, we also performed multiscale augmentation on each patch before model training to enhance scale invariance and simulate varying tissue magnifications. This approach may be particularly valuable in histopathology, where features of interest, such as nuclei and staining intensity, may appear at different scales across slides. We generated downscaled versions of each patch using four scale factors (1.0, 0.75, 0.5, and 0.25) and centred them within a canvas of original dimensions filled with the estimated background colour. This maintained spatial consistency and context while adding natural variations in patch scale, as shown in **Supplementary Figure S4**. Finally, we trained our regression model on 14244 (3561 x 4) patches; furthermore, we split this dataset into training (11396) and validation (2848) sets.

### Regression Model Architecture and Training

We utilised a ViT-based model from the distillation with no labels (DINOv2) family, specifically the ‘ViT-Base Patch14 Reg4’ variant.^34,35^ This model was pre-trained using self-supervised learning techniques on extensive image datasets, enabling it to learn robust features. The architecture features 86 million parameters with 115 million activations, a 14×14 patch size, and additional registration tokens that enhance localised and contextual understanding, making it particularly appropriate for histopathology image analysis. This architecture was selected based on its superior validation performance, indicated by the lowest root mean squared error (RMSE) during initial screening (**Supplementary Figure S5**). We also determined the optimal hyperparameters (batch size, optimiser, and loss function) based on validation loss over three epochs with different combinations of the parameters, as highlighted in **Supplementary Figure S6**. The top three optimisers were further evaluated using the whole training data. The optimal hyperparameters identified for the model include a batch size of 32, the Adam optimiser, and a Mean Squared Error (MSE) loss function. The maximum batch size of 32 was selected due to computational constraints, as the model requires input images to be 518 x 518 pixels.

For the training of our regression model, we adjusted the actual pre-trained model’s classification head with a custom regression head tailored for two regression outputs. This modification allowed the model to adapt pre-trained representations to the specific task of Ki-67 quantification from histological patches. The final head included fully connected layers to provide two outputs, presenting the counts of positive and negative nuclei. The input patches were resized to 518 × 518 pixels, and a comprehensive augmentation pipeline was applied at the batch level to ensure diverse transformations of the same image. This enhances the model’s ability to learn invariant representations, important for distillation-based training.^36^ This included random horizontal and vertical flipping, rotations of up to 土 20 degrees, 土 5% zoom, and brightness and contrast adjustments of up to 5%. Perspective warping was applied with a factor of 0.05, and affine transformations and lighting adjustments (with a 75% probability) were used to enhance the model’s robustness to morphological and staining variability inherent in histopathology. The data normalisation was performed using ImageNet mean and standard deviation.

Our regression model, using pre-trained weights, was initially fine-tuned for a maximum of 20 epochs at a learning rate of 2e-3 and a weight decay of 2e-3, with an early stopping function used to monitor the validation loss. The training stopped if no improvement exceeding a threshold of 0.1 was observed for three consecutive epochs. Subsequently, all layers of the model were unfrozen, and a learning rate finder was used to determine the optimal learning rate. The model was trained for an additional 20 epochs using the 1cycle policy.^37^ This process utilised discriminative learning rates ranging from 1e-6 to 1e-4 and callback functions to enhance training effectiveness and performance for accurately quantifying the Ki-67 index.

### Classification Model for Patch Gradability

For quality control, we constructed a patch-level classification model for assessing the gradability of the extracted patches from WSIs to ensure they are clinically relevant for estimating the Ki-67 index. The classification model was trained to classify patches into three classes: gradable IHC-stained patches, gradable H&E-stained patches, and ungradable patches. Ungradable patches typically included background areas, ink markings, artefacts, tissue folds, hair, bubbles or poorly stained patches. The model filtered out these non-informative patches, ensuring only clinically relevant patches passed to the regression model.

We collected 7122 gradable patches for IHC-staining, comprising 3561 patches derived from the SHIDC-B-Ki-67 and BCData datasets and 3561 randomly selected from the ACROBAT dataset. For gradable H&E-stained patches, we preferred 7122 patches from the TCGA-UT dataset, encompassing 32 types of cancer with diverse resolutions. Regarding ungradable patches, we again collected 7122 patches from the ACROBAT dataset without overlapping. The dataset was split into a training set of 80% (17093) and a validation set of 20% (4273). For external (unseen) testing, 399 patches were selected. We had 133 IHC-stained patches from the BCData testing dataset, and to address the class imbalance, we randomly selected 133 patches each from H&E-stained patches from the SICAPv2 dataset, and ungradable patches from the ACROBAT dataset. There was no overlap between the WSIs in the training, validation, and testing sets.

We implemented the classification model using the same architecture backbone as the one used in the regression model. Before training, all input images underwent stain normalisation using the Macenko method.^38^ The data augmentation and transformations were also performed using the same steps and parameters employed in the regression model. However, this model was fine-tuned for a maximum of 10 epochs with a learning rate of 2e-3 and a weight decay of 3e-3; callback functions were utilised to stop training if the validation loss did not decrease by at least 0.1 for the subsequent three epochs. After this initial phase, the model was unfrozen, and training continued using the 1cycle policy, with a discriminative learning rate that varied from 1e-6 to 1e-4 for an additional 10 epochs, employing the same callback functions.

### External Validation and Feature Correlation Analysis

We assessed the regression model’s ability to generalise on unseen data to quantify the Ki-67 counts for both positive and negative cells. The model determines the Ki-67 index, calculated as the ratio of Ki-67-positive cells to total cells (positive and negative) within each patch or aggregated across WSIs. Although the external datasets were provided without Ki-67 labels, the IHC4BC dataset contained image-derived features such as the average DAB intensity from IHC-stained patches, and the total nuclei count from corresponding H&E-stained patches. We evaluated the biological significance and consistency of the predicted Ki-67 indices by conducting correlation analyses with the available morphological features.

### Visual Interpretability

We visualised the attention map using activations from the model’s patch embedding layer to highlight areas of focus during inference. Each image tensor was passed through the model, and we extracted intermediate activations from the patch projection layer. After averaging and normalising these activations, we created smooth 2-dimensional attention maps. These maps were upsampled and overlaid on the input images, producing visual heatmaps highlighting the most critical areas to the model’s predictions. The colourised map combined the original image, creating visually interpretable heat maps that localised influential features.

### Cross-Stained Pseudo-Labelling Strategy

We employed a cross-stain pseudo-labelling technique utilising our trained regression model to quantify the Ki-67 index from IHC-stained patches. Initially, the model was used to predict Ki-67 counts from the IHC-stained patches from the IHC4BC dataset. These patches were paired with their corresponding H&E-stained patches. Subsequently, the predicted Ki-67 counts from the IHC patches and their associated H&E patches were used to train a new, cross-modality model to compute Ki-67 counts from H&E-stained histology images.

We collected 20656 paired IHC and H&E-stained patches from the IHC4BC dataset. These were randomly split into subsets: 80% for model development (13220 for training and 3304 for validation) and 20% for testing (4132 patches). This model quantifies Ki-67 counts from H&E-stained histological images, analysing morphological features in patches without requiring further IHC staining. Additionally, the cross-modality model was trained solely on the Ki-67 index, bypassing the counts of positive and negative cells.

### Slide-Level Ki-67 quantification and Hotspot detection

We developed a comprehensive pipeline to quantify Ki-67 expression at the slide level, including tissue detection, patch extraction, gradable patch filtering, Ki-67 count estimation, and hotspot highlighting within WSIs. Initially, WSIs were preprocessed for automated tissue detection at low resolution using the OpenSlide library^39^, applying grayscale thresholding and morphological operations to remove background artefacts. We filtered regions based on solidity (≥0.3), aspect ratio (0.1–10.0), and a minimum tissue area of 5000 pixels to retain only relevant tissue morphologies. Non-overlapping patches of 518 x 518 pixels were extracted, and each patch was analysed for quality metrics such as tissue coverage (≥2%), brightness consistency (0.03–0.97), edge density (≥2%), and brightness variation (≥0.02 standard deviation), excluding low-quality or irrelevant regions. Filtered patches underwent thorough quality control based on several criteria, such as tissue presence, brightness consistency, local texture variation, and edge sharpness. Accepted patches were padded using the estimated background colour from the outer edge pixels to ensure uniform input dimensions and mitigate border artefacts during model inference. Afterwards, we employed the classification model to distinguish gradable (IHC and H&E) patches from ungradable patches. We processed only the gradable patches for calculating the Ki-67 index using the models. Finally, we aggregated all the predicted values from each gradable patch from WSIs to quantify the total counts of positive and negative cells.

In addition to global Ki-67 quantification, we performed spatial analysis to identify high-proliferation regions (hotspots) within each WSIs. A patch-level heatmap illustrating the predicted fraction of Ki-67-positive cells was converted to a binary format using an optimal cut-off threshold to identify hotspot regions.^40^ For qualitative assessment, we created composite visualisations that included the original WSI thumbnails, a hotspot overlay, a bar chart displaying the counts of predicted positive and negative nuclei, and their corresponding Ki-67 index. These visualisations assist pathologists in timely analysing and evaluating tumour proliferation heterogeneity across different regions.

### Models Evaluation and Statistical Analysis

The models’ performance was evaluated against actual and predicted values. The performance of the regression models was evaluated using standard regression metrics, including the mean absolute error (MAE), RMSE, and the coefficient of determination (R²). For the classification task, predictions were assessed using multiple classification metrics, including accuracy, precision, recall, F1-score, and area under the receiver operating characteristic curve (AUROC) for each class compared to all other classes.^41^ The classification outputs were derived from softmax predictions, with the final class being determined by the maximum probability. Executing 4000 bootstrap iterations for each metric ensured statistical rigour with the estimation of 95% confidence intervals (CI).^42^

## RESULTS

### Regression Model Performance

The regression model effectively estimated the counts of positive and negative Ki-67-stained nuclei from histological images, underscoring the potential of a ViT-based regression model in computational pathology. The model demonstrated strong predictive performance on the validation set, achieving an MAE of 9.13 (8.86, 9.42), indicating the predicted counts deviated from the ground truth by an average of approximately nine cells per patch. The RMSE was 13.99 (13.43, 14.58), indicating a low average deviation from ground truth cell counts. Importantly, the model achieved an R² of 0.90 (0.89, 0.91) with a 95% CI, indicating that it accurately captured 90% of the variance in actual positive and negative cell counts.

Our regression model also demonstrated strong performance on the unseen testing set, as evidenced by the high R² value, which surpassed 0.90 for both positive and negative cell counts. In addition to the individual cell counts, the model demonstrated a strong ability to estimate the patch-level Ki-67 index, achieving an R² value of 0.93. All regression metrics are presented in **Table 2**, which includes the 95% CI for each class. **Figure 1 (A)** visualises the agreement between actual and predicted cell count values using Bland-Altman plots. These results highlight the model’s robustness and reliability in quantifying cellular proliferation from IHC-stained tissue patches.

**Table 2.** Regression model performance on our unseen testing dataset.

| Class | MAE (95% CI) | RMSE (95% CI) | R<sup>2</sup> (95% CI) |
| --- | --- | --- | --- |
| Positive Cells | 8.07 (6.41,10.01 ) | 13.12 (8.87, 17.54) | 0.92 (0.90, 0.94) |
| Negative Cells | 16.02 (13.55, 18.56) | 21.66 (18.05, 25.50) | 0.90 (0.85, 0.94) |
| Ki-67 Index | 4.39 (3.70, 5.15 ) | 6.15 (5.21, 7.08) | 0.93 (0.91,0.95) |

**Abbreviations:** MAE: Mean Absolute Error; RMSE: Root Mean Squared Error; R<sup>2</sup>: Coefficient of Determination; CI: Confidence Intervals.

**Figure 1.**
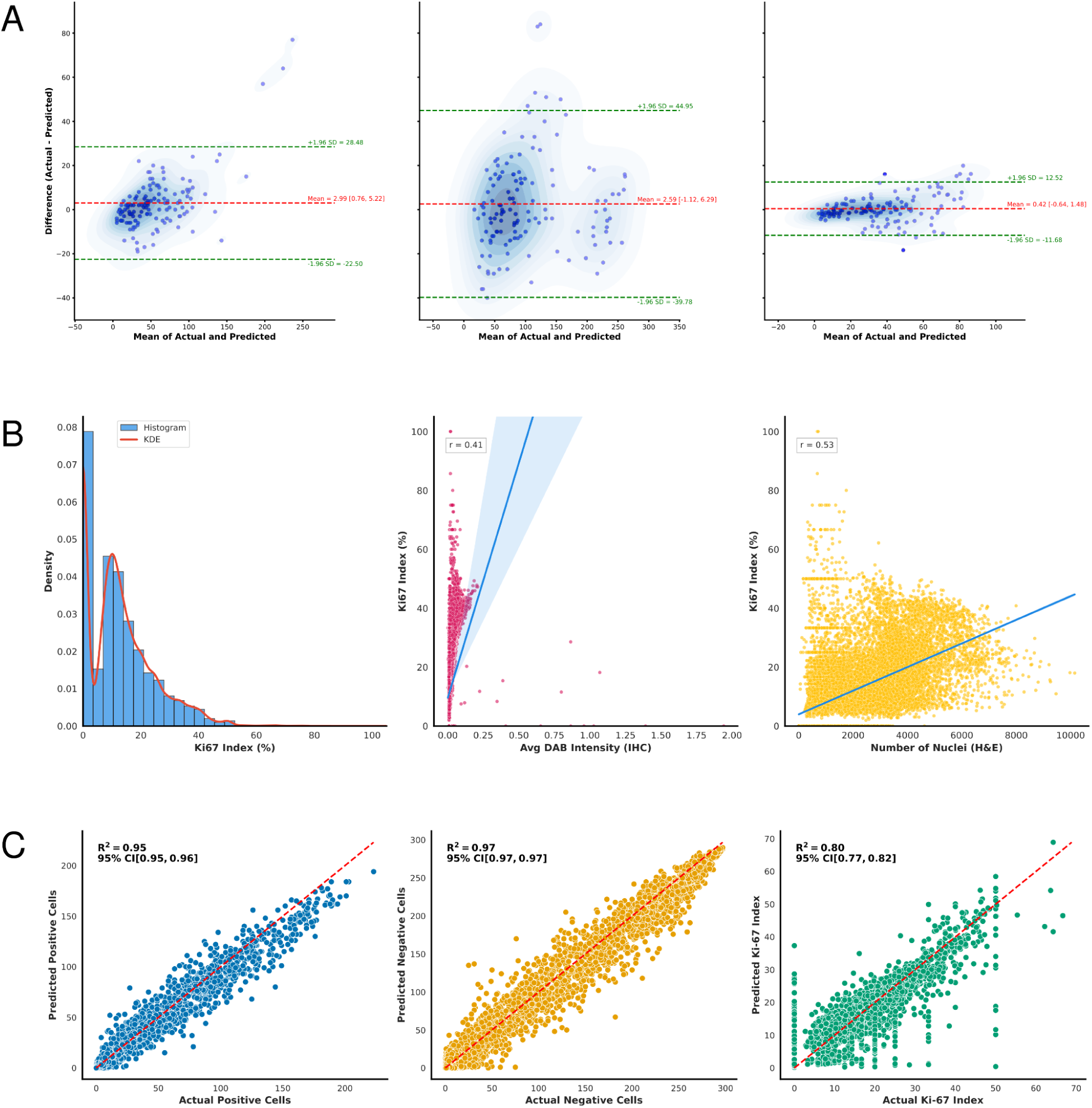
Patch-level agreement and morphologic correlation for Ki-67 quantification using IHC regression model. (**A**) Bland-Altman plots show the agreement between actual and predicted cell count values with the Ki-67 index in the testing set, highlighting mean differences and 95% limits of agreement. The left subplot displays positive counts, the middle subplot indicates negative counts, and the right subplot exhibits the Ki-67 index. (**B**) The IHC4BC dataset indicates a right-tailed distribution of the predicted Ki-67 index, with moderate to strong positive correlations (correlation coefficient = r) to DAB intensity and total nuclei count. (**C**) The cross-modality regression model quantified the Ki-67 counts on the testing set (4132), visualising the relationship between actual and estimated cell counts for positive, negative, and Ki-67 index.

### Classification Model Performance

The classification model obtained superior performance in distinguishing between H&E-stained, IHC-stained, and ungradable patches. On both the validation and unseen testing sets, the model achieved an overall accuracy of 100%. **Supplementary Tables S1 and S2** present the diverse classification metrics for each class on the validation and testing sets, respectively. This classification model was essential in filtering out low-quality patches, including those affected by artefacts, ink markings, and tissue folds, to ensure that clinically relevant IHC-stained patches were analysed for quantifying the Ki-67 index from WSIs.

### External Validation and Morphological Feature Associations

The regression model estimated the Ki-67 cell counts on DeepSlide and IHC4BC datasets to assess its generalisability and biological plausibility. In the DeepSlide dataset, predicted cell counts were visualised in **Supplementary Figure S7**. Despite the absence of actual labels, the model successfully inferred biologically meaningful proliferation patterns. Most values clustered below 25% of the Ki-67 index, reflecting the expected heterogeneity in tumour proliferation across tissue regions. The structured pattern of predictions reflects consistency between the model’s outputs, including both positive and negative counts. Similarly, in the IHC4BC dataset, scatterplots of predicted positive versus negative nuclei count highlighted patches with exceptionally high Ki-67 index values—some reaching 75%, 85%, and even 100% (**Supplementary Figure S8**). The predicted values highlighted the typical heterogeneity of breast cancer tissues and inferred biologically relevant proliferation patterns, even without annotated labels.

Further interpretation of the predictions and exploration of their biological relevance involved analysing correlations between the predicted Ki-67 index and two morphological features available in the external datasets: average DAB intensity (from IHC-stained patches) and total nuclei count (from H&E-stained patches). The result indicated a moderate positive correlation between the Ki-67 index and average DAB intensity, suggesting that regions with stronger chromogenic signals exhibit higher proliferative activity. Additionally, a stronger correlation (correlation coefficient = 0.53) was found between the Ki-67 index and total nuclei count, suggesting areas with higher cellular density are more likely to correspond with increased predicted proliferation, as illustrated in **Figure 1 (B).** The associations between the predicted Ki-67 index and tissue morphology suggest considerable potential for utilising this model in exploratory analyses, automated reporting, and clinical utility where manual annotations are unavailable.

### Clinical Transparency with Attention Maps

The attention maps explained the decision-making processes of both models in a clinical context. The regression model emphasised regions with dense nuclear content, particularly in areas predicted to have high Ki-67-positive counts. These regions align closely with histologically significant tumour areas exhibiting increased proliferative activity. The attention distribution was more continuous and diffuse, capturing subtle spatial variations in Ki-67 expression throughout the tissue, as shown in **Figure 2**. In contrast, the classification model generated more defined and localised attention maps, as illustrated in **Figure 3**. For gradable IHC and H&E patches, the model accurately focused on cellular areas with apparent morphological features, while for ungradable patches, the attention maps were diffuse or focused on the background, indicating minimal diagnostic content. Our models demonstrated transparent and biologically relevant attention patterns, confirming their reliable predictions and potential utility in computational pathology.

**Figure 2.**
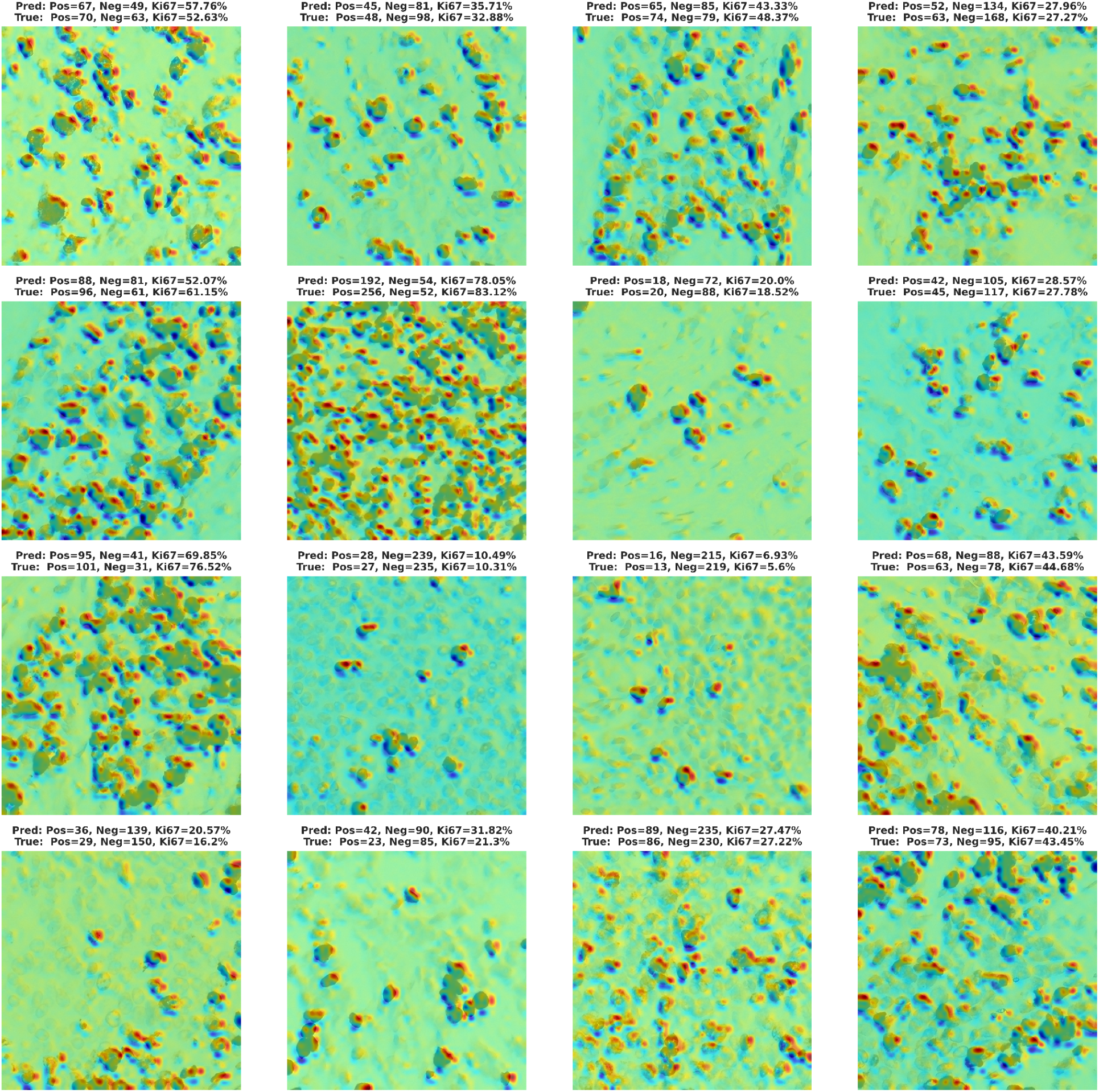
Our model predicted Ki-67 cell counts against the actual label from the testing set. The heatmap overlay displays the model-predicted distribution of Ki-67-positive cell counts, with warmer colours indicating a higher predicted Ki-67 index, aligning with ground truth.

**Figure 3.**
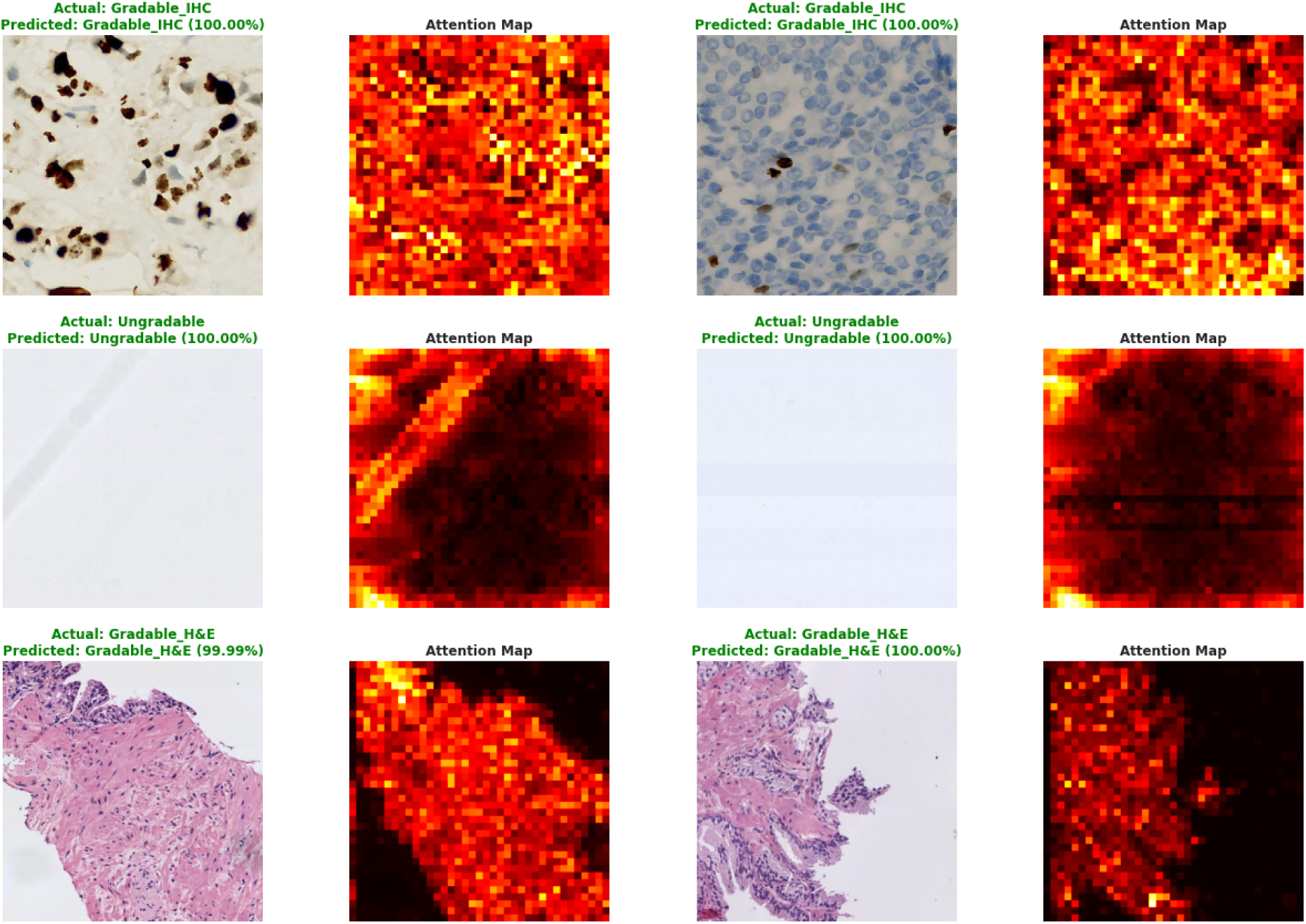
The classification model was evaluated on randomly selected patches from the testing set, which included gradable H&E, gradable IHC, and ungradable patches. The model’s class predictions with confidence levels were compared to the ground truth labels. Attention maps identify critical areas that influence the model’s decisions, highlighted by brighter regions.

### Cross-Modality Model Performance

The cross-modality (H&E regression) model demonstrated robust performance in quantifying both positive and negative cell counts, achieving overall MAE and R² values of 7.28 (7.04, 7.54) and 0.97 (0.97, 0.97) with 95% CI, respectively, on the validation set. The model also indicated strong consistency on the testing set (**Supplementary Table S3**), obtaining high agreement for positive (R²: 0.95) and negative (R²: 0.97) cell estimations. **Figure 4** exhibits paired patches for visual assessment of IHC-driven (model-inferred) labels and cross-modality model estimations on H&E-stained patches. The cross-modality model achieved an R² value of 0.82 (0.79, 0.84) on the validation set and 0.80 (0.78, 0.82) on the testing set when trained solely on the Ki-67 index, demonstrating its ability to learn visual patterns of cellular proliferation directly from histological patterns without relying on cell counts, as depicted in **Figure 1(C)**. The model accounted for approximately 80% of the variance in Ki-67 index values derived from corresponding IHC-stained patches. The random paired patches were selected from the testing set to visualise the patches with IHC-driven labelled and predicted Ki-67 indices from H&E-stained patches, which are visualised in **Supplementary Figure S9**.

**Figure 4.**
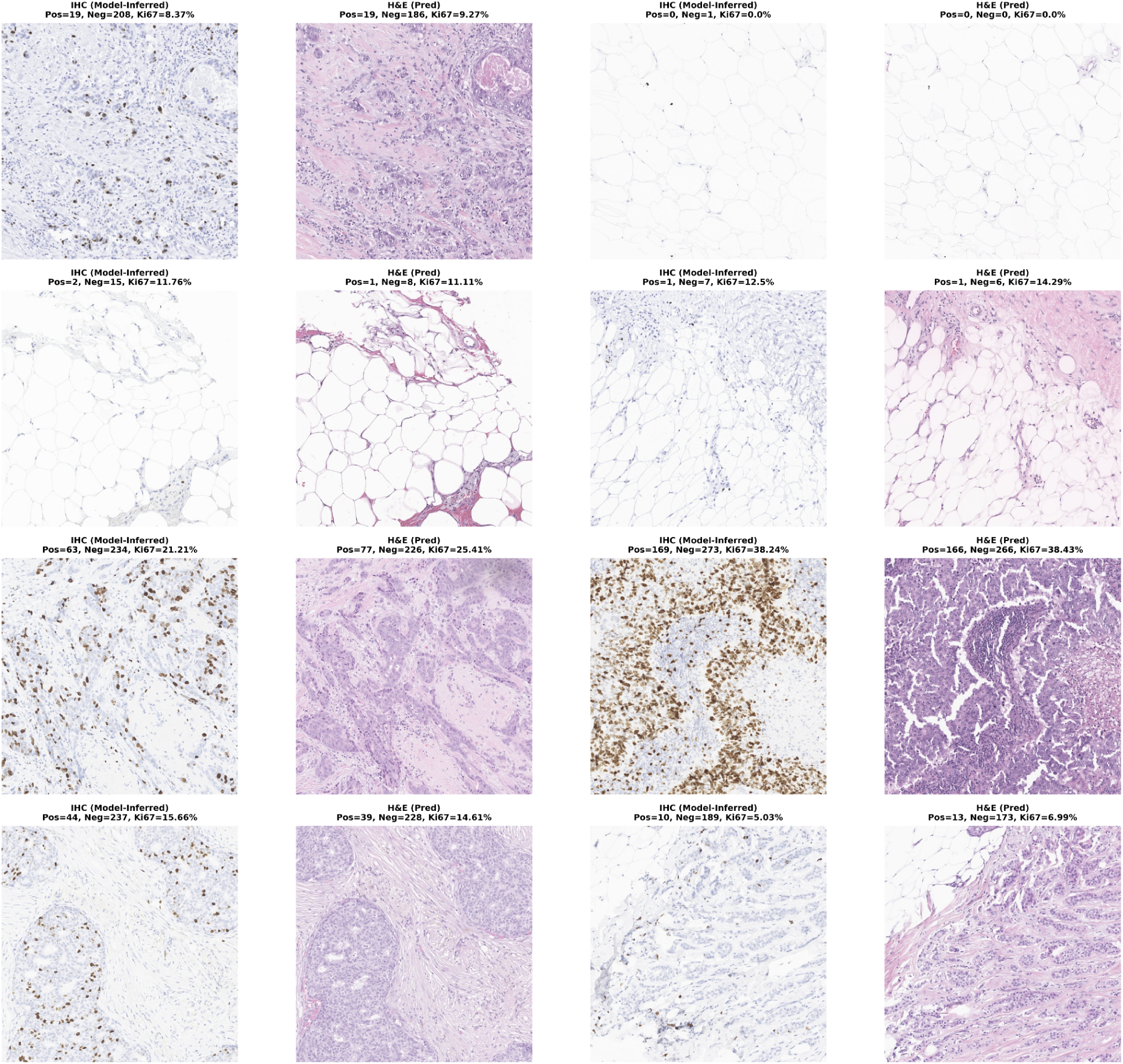
Visualisation of Ki-67 expression from paired IHC and H&E-stained patches is illustrated as follows: (IHC → H&E) | (IHC → H&E). The IHC patches indicate Ki-67 positive and negative cell counts inferred from the IHC regression model, while the corresponding H&E patches display Ki-67 cell counts quantified by the cross-modality model for the same tissue region across different staining modalities.

### Slide-Level Ki-67 Quantification and Hotspot Detection

Our trained models automatically quantified the Ki-67 index and hotspot detection from the ACROBAT dataset, containing 1996 unannotated IHC and H&E-stained WSIs. All the WSIs were used to determine Ki-67 cell counts, underscoring the positive correlation between predicted positive and negative counts. As expected, most IHC-stained WSIs clustered in the lower count ranges illustrated in **Supplementary Figure S10**. The Ki-67 index varied widely across the IHC-stained slides, capturing biological variability in tumour proliferation. We selected four WSIS covering a spectrum of predicted Ki-67 indexes—low, moderate, and high proliferation levels—to assess model performance and interpretability qualitatively. We visualised the corresponding hotspot region for each selected slide by identifying the most proliferative patch cluster, which was defined as the area with the highest fraction of Ki-67-positive cells. Additionally, the Ki-67 index quantification and hotspot highlighting from IHC-stained WSIS (843) are depicted in **Supplementary Figure S11**, utilising the IHC regression model. **Figure 5** depicts the distribution of predicted positive and negative cell counts from H&E-stained WSIs with hotspots using the cross-modality model. Our method provides a scalable and interpretable solution for automated Ki-67 analysis in real-world computational pathology workflows by quantifying Ki-67 expression at the slide level through histologically meaningful hotspot detection from both H&E and IHC-stained WSIs.

**Figure 5.**
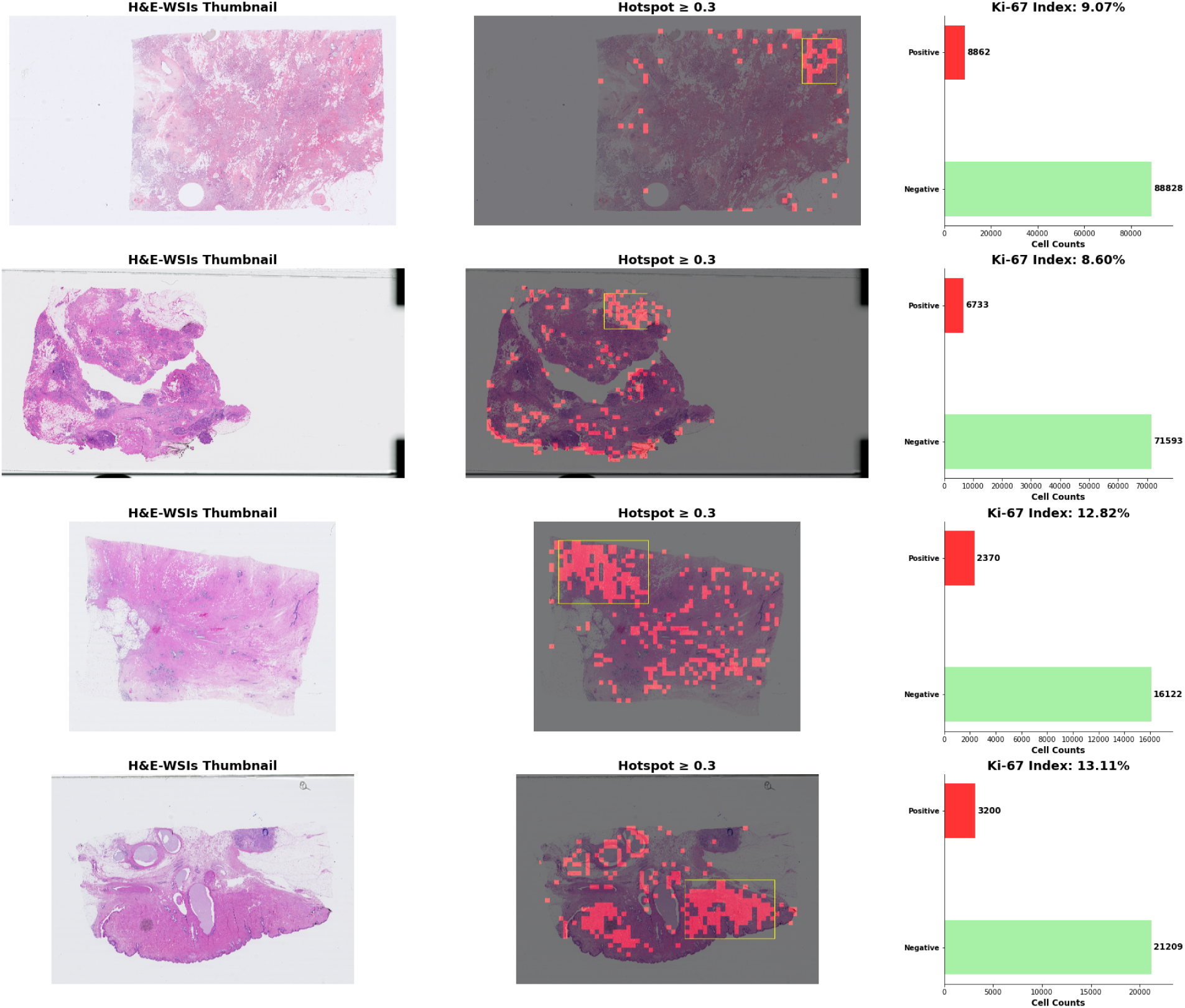
Quantification of Ki-67 index at slide-level and visualisation of hotspot using H&E-stained WSIs from the ACROBAT dataset. For each slide, the left side displays the original thumbnail of WSIs, the middle panel overlays predicted Ki-67 hotspots in red, with the largest hotspot region outlined (yellow), and the right panel presents bar plots of predicted Ki-67-positive (red) and Ki-67-negative (green) cell counts.

## DISCUSSION

Digital image analysis (DIA) tools hold the potential for automating Ki-67 scoring in breast cancer, but currently, none meet the clinical standard for reliability, although a deep learning-based algorithm shows promise in quantifying the Ki-67 index.^18,43,44^ Our study introduces a comprehensive pipeline using the backbone of the pre-trained ViT-based architecture for developing a regression model for quantifying the Ki-67 index from WSIs in breast cancer histopathology. The model’s global self-attention mechanism facilitates the effective modelling of long-range dependencies and contextual relationships among nuclei. It is particularly advantageous in densely packed or overlapping cellular regions where traditional segmentation or detection models often underperform due to their limited receptive fields and reliance on localised spatial features.^45,46^ This approach effectively identifies and quantifies Ki-67 cell counts by integrating quality control steps with the regression model, significantly enhancing potential utility in digital pathology workflows.

The regression model can automatically learn to determine a specific threshold (cell detection cut-off threshold) for distinguishing between positive (brown) and negative (blue) nuclei, which usually varies substantially among pathologists.^47^ The model accurately estimated positive and negative Ki-67 cell counts by automatically setting a threshold for stained cells, achieving high R² values (**Table 2**) on the testing set. The model’s precision, demonstrated by low MAE, highlighted that the predicted positive and negative counts deviated from the ground truth by an average of approximately eight and 16 cells per patch, respectively. In the training dataset, the mean counts for positive and negative cells were 30 and 60, as illustrated in **Supplementary Figure S3**. This underscores its potential for automated and consistent assessment of tumour proliferation, addressing the variability in manual scoring often encountered in anatomical pathology practice.

In addition to improving the robustness of our regression model for Ki-67 quantification, we implemented a multi-scale augmentation strategy during model training to enhance scale invariance and simulate varying tissue magnifications. We generated downscaled versions using four scale factors, allowing the model to capture both local and global morphological features across various resolutions. While most studies in histopathology have focused on segmentation or classification tasks, similar multi-scale strategies were applied in other imaging domains. For instance, Yang et al. developed an automatic analysis framework based on 3D-CT multi-scale features to predict Ki-67 expression levels in renal cell carcinoma, demonstrating that multi-scale feature extraction significantly improves prediction performance.^48^ While their study focused on CT images, the core idea—training with multi-scale data improves model generalisation. Thus, training with multi-scale augmented data enhances the accuracy and reproducibility of Ki-67 quantification by mitigating field-of-view bias and promoting generalisability across diverse datasets. For example, the IHC4BC data display patches at a lower magnification, whereas our model identifies cell counts based on clinically relevant morphology for both positively and negatively stained cells, which are highlighted with attention maps, as shown in **Supplementary Figure S11**.

The regression model objectively quantified the positive and negative cells at the slide level from the ACROBAT dataset, reflecting spatial heterogeneity throughout the entire tumour bed. Our approach addresses the limitations associated with manual hotspot selection and field-of-view bias.^43^ Notably, most tumours in the dataset exhibited a Ki-67 index below 20% (**Supplementary Figure S10**), aligning with a lower proliferative profile commonly associated with luminal A-like subtypes.^49^ This directly implies clinical decision-making in surgical oncology, where Ki-67 serves as a crucial biomarker for post-operative risk stratification and determining the necessity of chemotherapy. In current practice, variability in manual scoring can lead to inconsistent treatment opinions, particularly in borderline cases (comprising 15% to 25% of the Ki-67 index). We provide robust and reproducible quantification of the Ki-67 index at the slide level, reducing observer bias and facilitating personalised treatment. Additionally, quantifying cell counts and generating hotspots in case-level reports enhances pathology workflows, potentially providing a decision-support tool in computational pathology. With prospective validation and clinical deployment, this approach could improve diagnostic accuracy, optimise therapeutic pathways, and ultimately enhance patient outcomes in breast cancer care.

Finally, we evaluated the generalisability of our regression model across different staining modalities using paired IHC and H&E-stained patches from the IHC4BC dataset (20656), comparing prediction correlation for Ki-67 counts. The results demonstrated substantial variability in model performance across stain modalities (**Supplementary Figure S13)**. The predictions for the counts of negative cells achieved an R² of 0.85, indicating a strong association between the model’s predictions based on IHC and H&E patches. However, the negative R² value (-46.40) for positive cell counts indicates the model’s predictions fail to generalise across stains. This evaluation across different modalities highlights the challenges in transferring our models between various staining protocols. While H&E slides provide valuable information about tissue morphology, the model fails to accurately capture molecular expressions, as revealed by IHC, which limits its ability to reflect proliferation markers. Thus, we adopted a pseudo-labelling strategy to train a regression model on H&E patches to overcome variability across stain modalities. The IHC-based regression model quantified Ki-67 counts from IHC-stained patches, which were used as surrogate labels for the corresponding H&E-stained patches. This method enabled the cross-modality H&E regression model to learn stain-specific morphological patterns, resulting in high accuracy in predicting both positive and negative cell counts. By optimising for the final Ki-67 index instead of intermediate cell counts, the model learns more abstract features that strongly associate (R² = 0.8) with the IHC-driven Ki-67 index. This accuracy is considerable, considering the subjectivity of manual scoring and the variability in staining, reducing the need for labour-intensive cell-level annotations while still providing clinically relevant results from H&E-stained slides. Thus, the cross-modality model demonstrates a viable option for calculating Ki-67 expression from standard H&E-stained histology images, thereby reducing dependence on IHC-staining and enhancing clinical utility in computational pathology.

This research has some important caveats and limitations. Although extensive validation was performed, prospective clinical validation on larger, multicenter datasets would provide more substantial evidence for clinical implementation. Second, the current model focuses solely on Ki-67 quantification; integration with additional biomarkers or clinical parameters could enhance predictive accuracy and clinical relevance. While multi-scale augmentation enhanced our model’s ability to capture features at various resolutions, it may not fully account for all real-world variations. Lastly, the cross-modality model needs validation against ground truth labels at WSIs, specifically with H&E-stained WSIs, which are not publicly available at this time. Future studies should focus more on extensive, multicenter, and diverse datasets, incorporating additional histological biomarkers.

In conclusion, our regression model accurately quantifies Ki-67 expression from IHC-stained WSIs in breast cancer. The cross-stained model also accurately projected molecular expression from morphological information, providing a reliable tool for estimating the Ki-67 proliferation index from standard H&E-stained slides without requiring additional IHC staining. Attention maps from the models demonstrated transparent and biologically relevant patterns, highlighting their potential utility in computational pathology. Further validation with extensively annotated WSIs datasets remains necessary. Future studies will focus on enhancing training datasets with diverse ethnicities and different scanners to develop a robust and generalised model for other cancer types.

## Data Availability

All the data utilised in this study are publicly available.

## ACKNOWLEDGEMENTS

This work was supported by an Australian National Health and Medical Research Council Leadership Award (A.W.H.).

