## Supplemental information for "Automated quantification of Ki-67 expression in breast cancer from H&E-stained slides using a transformer-based regression model"

**Affiliations:**
1. Pandani Solutions Pty Ltd, Hobart, Australia.
2. Menzies Institute for Medical Research, University of Tasmania, Australia.
3. Department of Pathology, Royal Hobart Hospital, Hobart, Australia.
4. School of Medicine, University of Tasmania, Australia.

***Correspondence:**

Abadh K Chaurasia

Menzies Institute for Medical Research,

University of Tasmania, Australia.

### SUPPLEMENTARY METHODS:

#### SHIDC-B-Ki-67

This dataset comprises 1656 training and 701 testing patches, each with cell coordinates for Ki-67-positive, Ki-67-negative, and Lymphocytes. The cell coordinates with patches are visualised in **Supplementary Figure S1**. Each patch contains an average of 69 cells, totalling 162,998 cells across the dataset. All the patients were included with a pathologically confirmed diagnosis of breast cancer at Shiraz University of Medical Sciences, Pathology Laboratories, Iran.[1] It contains microscopic tru-cut biopsy images of malignant breast tumours, exclusively of the invasive ductal carcinoma type.[2] The Olympus BX-51 system microscope with a relay lens was coupled to an OMAX digital colour camera A35180U3 to obtain digital images of the tumour tissue slides.

#### Breast Tumour Cell Dataset

The Breast Tumour Cell Dataset (BCData) includes 1338 patches extracted from 394 WSIs, containing 181074 labelled tumour cells, 62623 positive and 118451 negative cells.[3] The WSIs were scanned with a Motic BA600-4 scanner at 40x magnification (0.2239 μm/pixel), and all data were anonymised. The evaluation of positive and negative expressions of Ki-67 was performed by a team of ten expert pathologists, utilising a rigorous three-stage labelling process. This process included the initial classification of tumour cells, a systematic review, and final validation by a chief expert pathologist. We highlight the annotated patches in Supplementary Figure S2 to demonstrate the diversity and complexity of cellular patterns. Furthermore, the dataset was divided into training (52%), validation (12%), and testing (36%) subsets.

In our study, we trained a regression model using the SHIDC-B-Ki-67 and BCData datasets, keeping BCData's validation set separate for testing due to its accurate labelling by an expert team of pathologists. We enhanced model performance by combining both datasets and randomly splitting them in an 80:20 ratio, resulting in 2,849 patches for training and 712 for validation, while reserving 12% of BCData for testing. **Supplementary Figure S3** illustrates the distribution of cell counts used to train the regression model across both datasets.

**DeepSlides**

It contains 694 patches (512 x 512 pixels) from WSIs of 32 breast cancer patients, without disclosing the labelled Ki-67 positive and negative stained cells or their coordinates.[4] The slides were scanned using the Aperio ScanScope at a magnification of 40x (0.2461 × 0.2461 $\mu m^2$). The tumour and non-tumour regions on the slides were identified and marked by an experienced breast pathologist. Our study used this data to determine the Ki-67 index and visualise the distribution of predicted cell counts across total samples, generating quantitative insights into Ki-67 expression patterns.

**IHC4BC**

The immunohistochemistry for Breast Cancer (IHC4BC) dataset includes comprehensive histopathology patches designed for predicting key breast cancer biomarkers, particularly estrogen receptor, progesterone receptor, and Ki-67 from Hematoxylin and Eosin (H&E) and IHC-stained breast cancer tissue images.[5] It comprises approximately 90,000 pairs of H&E and IHC-stained images, with corresponding CSV files containing 3,3'-diaminobenzidine (DAB) intensity measurements per nucleus and total nuclei counting in the H&E patches.

Our study exclusively utilised H&E- and IHC-stained paired patches (20656) to validate the regression model. Although the dataset lacks direct information regarding the ground truth for the Ki-67 index, the intensity of the DAB staining can be used to indirectly assess the predicted proliferation index and explore the relationship between the predicted Ki-67 index and DAB intensity and cell counts extracted from the corresponding H&E-stained patches.

**ACROBAT**

The AutomatiC Registration Of Breast cAncer Tissue (ACROBAT) dataset consists of 4212 WSIs from 1153 female patients diagnosed with primary breast cancer.[6] Each patient has one H&E-stained slide and up to four additional slides stained with IHC targeting estrogen receptor, progesterone receptor, human epidermal growth factor receptor, and Ki-67. The slides were digitised using Hamamatsu NanoZoomer scanners at a magnification of 10x, with a resolution of approximately 0.92 micrometres per pixel, providing multiple resolution levels. The dataset was divided into training (3406 WSIs from 750 patients), validation (200 WSIs from 100 patients), and test subsets (606 WSIs from 303 patients). We collected 843 IHC-stained WSIs to highlight the hotspot and quantify the Ki-67 index at the slide level.

This dataset generated gradable and ungradable IHC-stained patches from 40 randomly selected WSIs, splitting equally for training and testing. We manually evaluated and classified the patches into gradable and ungradable classes (blurred tissue, artefacts, ink, folds, bubbles, and hair). We ultimately selected 3561 gradable patches and 7122 ungradable patches to develop a classification model to assess the quantity of extracted patches from the WSIs. Additionally, we selected 133 ungradable patches to test the classification model. These ratios were decided to ensure a balanced representation among the three classes.

**TCGA-UT**

The Cancer Genome Atlas - Uniform Tumour (TCGA-UT) dataset contains 1608060 patches, each measuring 256 × 256 pixels.[7] These patches were extracted from H&E-stained WSIs, representing 32 types of solid tumours across 7175 patients. The patches were selected from diagnostic slides within The Cancer Genome Atlas (TCGA) database, ensuring a diverse representation of tumour morphology while maintaining high-quality standards; 926 low-quality slides were removed. This dataset includes multi-resolution patches captured at various micrometres (μm) per pixel scale, ranging from 0.5 μm/pixel to 1.0 μm/pixel.

This dataset was explicitly utilised to train our classification model for quantity control, distinguishing between H&E-stained gradable, IHC-stained gradable, and ungradable patches. We randomly selected 3561 patches from all types of cancer with diverse resolutions, ensuring class balance by choosing an equal number of patches from each class with diverse resolutions.

**SICAPv2**

The SICAPv2 dataset is a publicly accessible collection of H&E-stained patches for prostate cancer.[8] The dataset includes 155 prostate biopsy samples collected from 95 patients, with each slide reviewed and labelled by expert urogenital pathologists. The WSIs were scanned at 40x magnification. We utilise this dataset to test our classification model externally.

### SUPPLEMENTARY FIGURES AND TABLES:

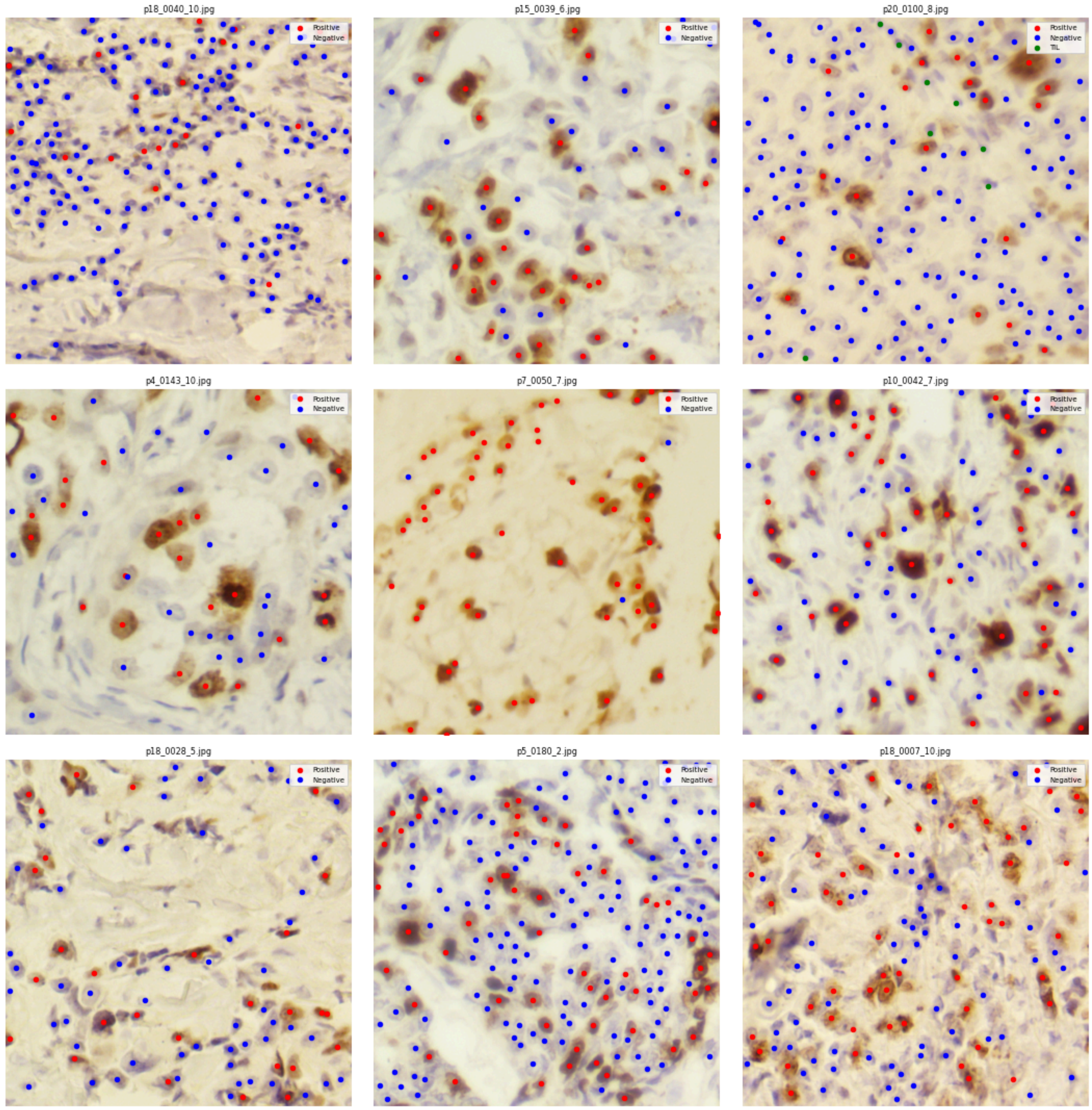


**Supplementary Figure S1.** *Randomly selected patches from the SHIDC-B-Ki-67 dataset with annotated coordinates; Positive Ki-67 cells are highlighted in red, Negative cells in blue, and Tumour Infiltrating Lymphocytes (TILs) in green.*

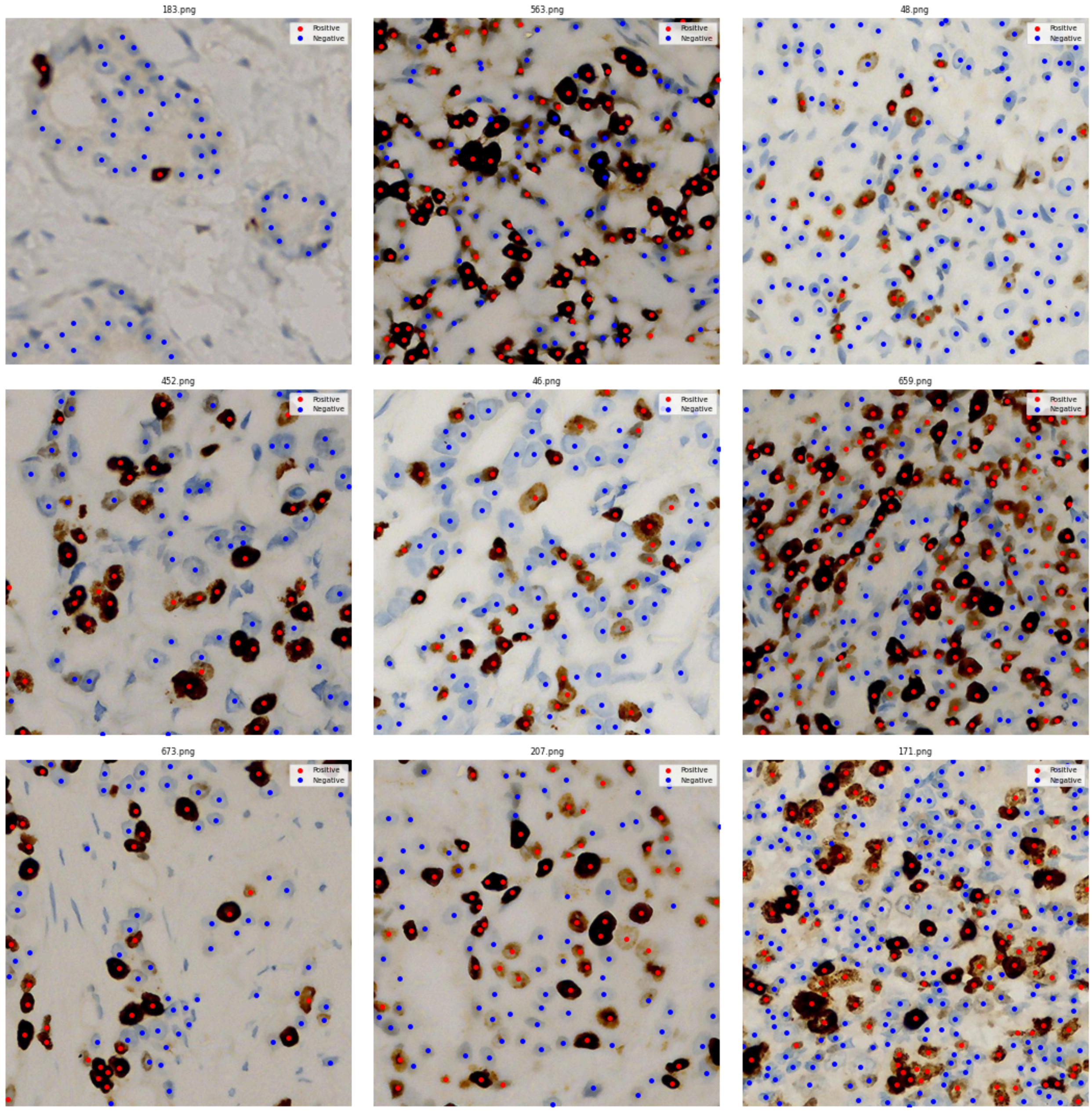


**Supplementary Figure S2.** *Randomly selected patches from the BCData set were visualised, with red dots representing Ki-67 positive cells and blue dots for Ki-67 negative cells, extracted from the corresponding annotation files.*

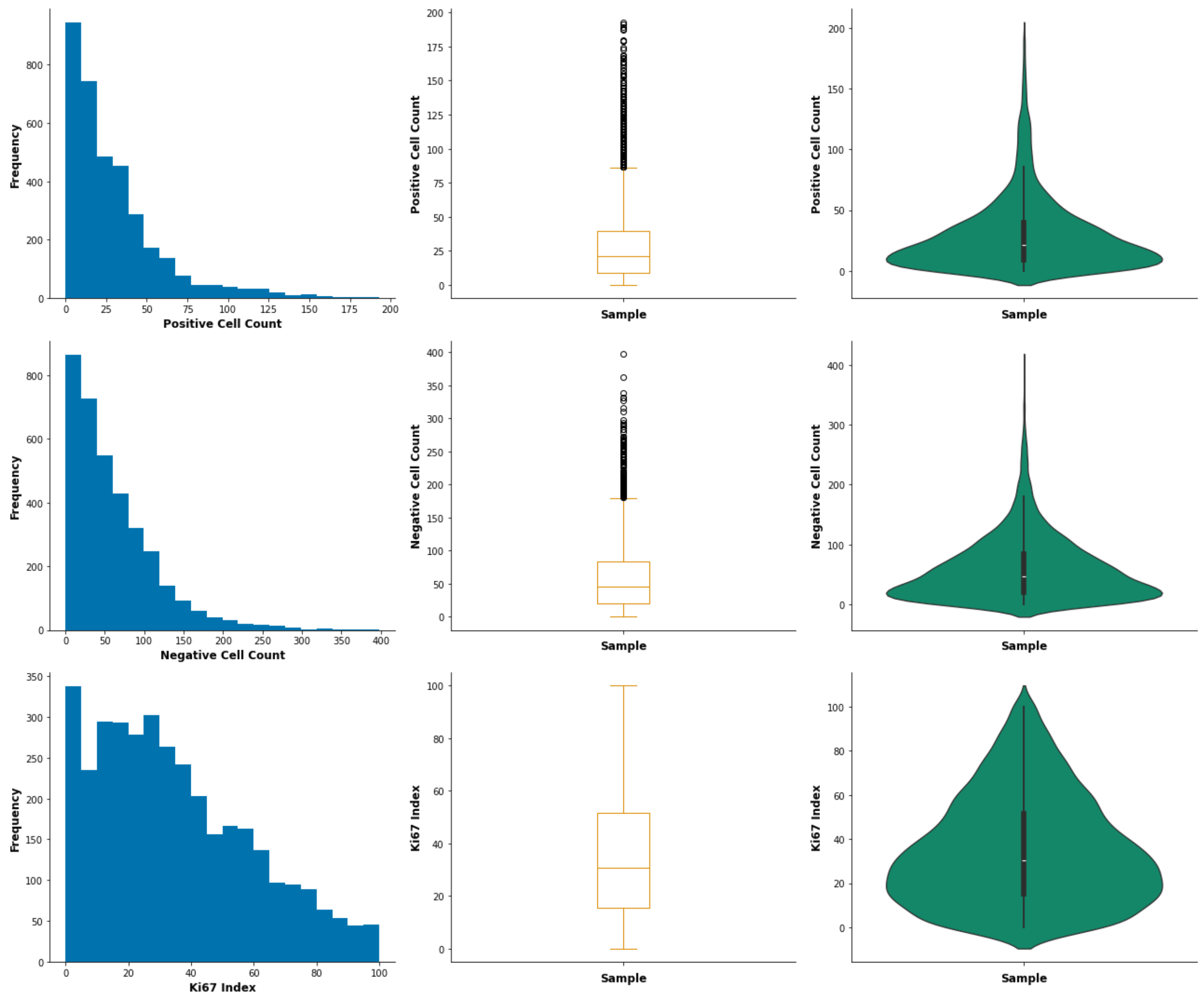


**Supplementary Figure S3.** *Distribution of positive, negative cell counts, and the Ki-67 index used for training the regression model from the SHIDC-B-Ki-67 and BCData datasets. The first column displays histograms showing the frequency distribution of each variable. The second column shows box plots summarising the median, interquartile range, and outliers. The third column presents violin plots depicting the kernel density estimation and distribution shape for positive cell counts, negative cell counts, and the Ki-67 index.*

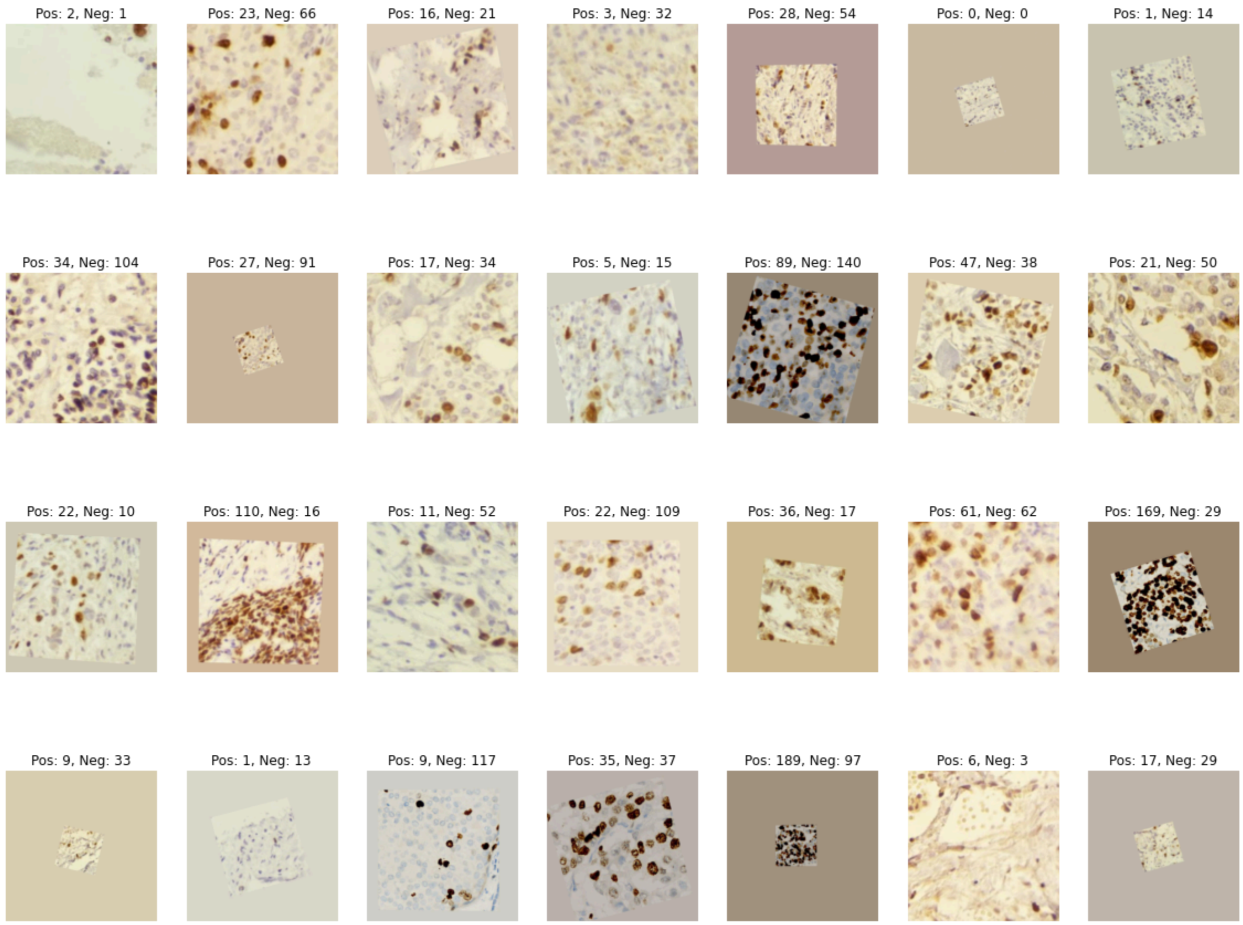


**Supplementary Figure S4.** *Random patches were selected from the training set, labelling positive and negative cell counts from IHC-stained patches.*

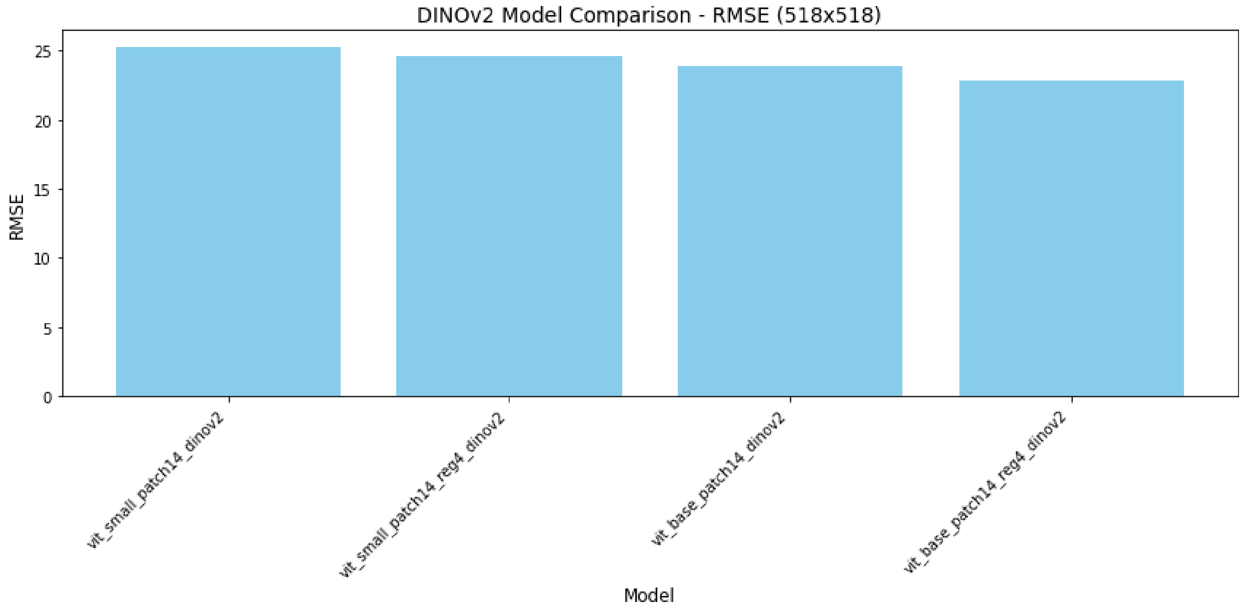


**Supplementary Figure S5.** *The optimal model was selected from variants of vision transformer architectures evaluated on 3561 images to quantify Ki-67 counts from IHC-stained patches, based on validation loss after three epochs.*

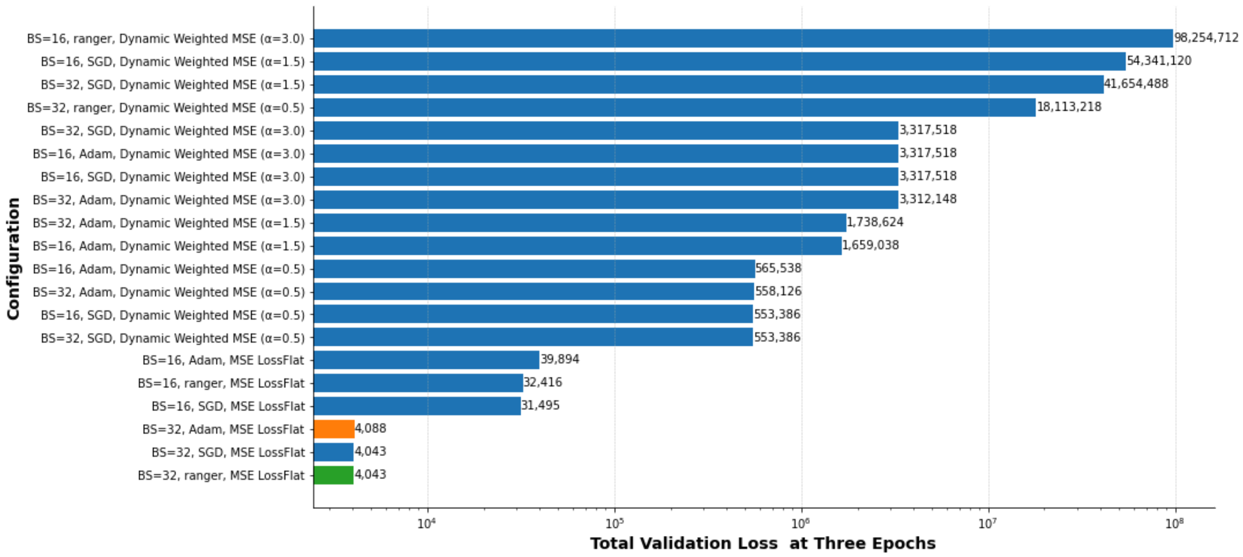


**Supplementary Figure S6.** *The different combinations of hyperparameters (batch size, optimiser, and loss function) were utilised to find the optimal set for accurately estimating the Ki-67 index from the total training dataset (11396).*

**Supplementary Table S1.** *Performance of the classification model on the validation set.*

| Class | n | AUROC Mean [95% CI] | Accuracy Mean [95% CI] | Precision Mean [95% CI] | Recall Mean [95% CI] | F1-score Mean [95% CI] |
|---|---|---|---|---|---|---|
| **Gradable_H&E** | 1454 | 1.000 [1.000, 1.000] | 1.000 [1.000, 1.000] | 1.000 [1.000, 1.000] | 1.000 [1.000, 1.000] | 1.000 [1.000, 1.000] |
| **Gradable_IHC** | 1411 | 1.000 [1.000, 1.000] | 0.999 [0.998, 1.000] | 0.999 [0.996, 1.000] | 0.998 [0.995, 1.000] | 0.998 [0.997, 1.000] |
| **Ungradable** | 1408 | 1.000 [1.000, 1.000] | 0.999 [0.998, 1.000] | 0.998 [0.995, 1.000] | 0.999 [0.996, 1.000] | 0.998 [0.997, 1.000] |

**Abbreviations:** **n** = Total number of patches; **AUROC**: Area Under the Receiver Operating Characteristic curve; **CI**: Confidence Intervals

**Supplementary Table S2.** *Performance of the classification model on the testing set.*

| Class | n | AUROC Mean [95% CI] | Accuracy Mean [95% CI] | Precision Mean [95% CI] | Recall Mean [95% CI] | F1-score Mean [95% CI] |
|---|---|---|---|---|---|---|
| **Gradable_H&E** | 133 | 1.000 [1.000, 1.000] | 1.000 [1.000, 1.000] | 1.000 [1.000, 1.000] | 1.000 [1.000, 1.000] | 1.000 [1.000, 1.000] |
| **Gradable_IHC** | 133 | 1.000 [1.000, 1.000] | 1.000 [1.000, 1.000] | 1.000 [1.000, 1.000] | 1.000 [1.000, 1.000] | 1.000 [1.000, 1.000] |
| **Ungradable** | 133 | 1.000 [1.000, 1.000] | 1.000 [1.000, 1.000] | 1.000 [1.000, 1.000] | 1.000 [1.000, 1.000] | 1.000 [1.000, 1.000] |

**Abbreviations:** **n** = Total number of patches; **AUROC**: Area Under the Receiver Operating Characteristic curve; **CI**: Confidence Intervals

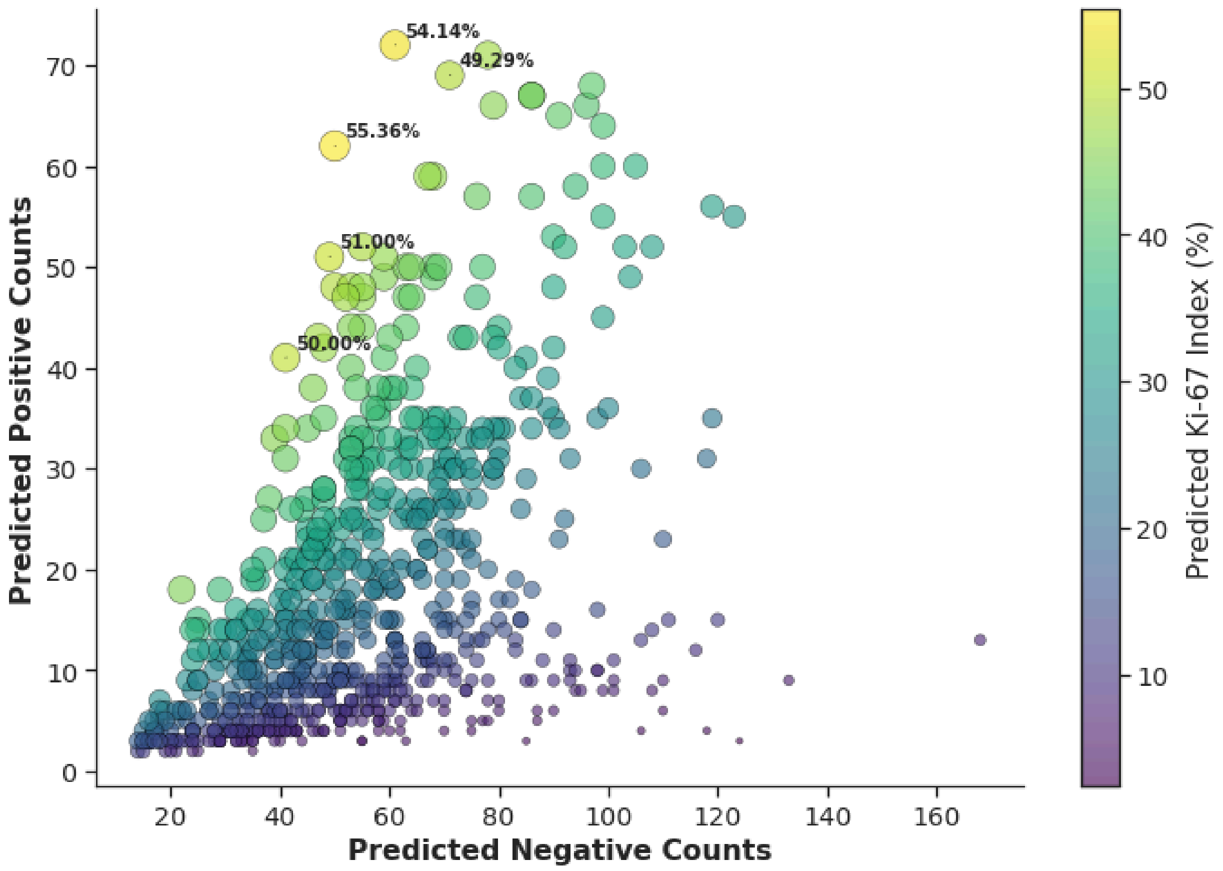


**Supplementary Figure S7.** *Scatter plot showing the relationship between predicted Ki-67 positive and negative cell counts across all the IHC-stained patches from the DeepSlide dataset. The size of each point represents the total number of predicted cells (both positive and negative), while the colour indicates the predicted Ki-67 index (%).*

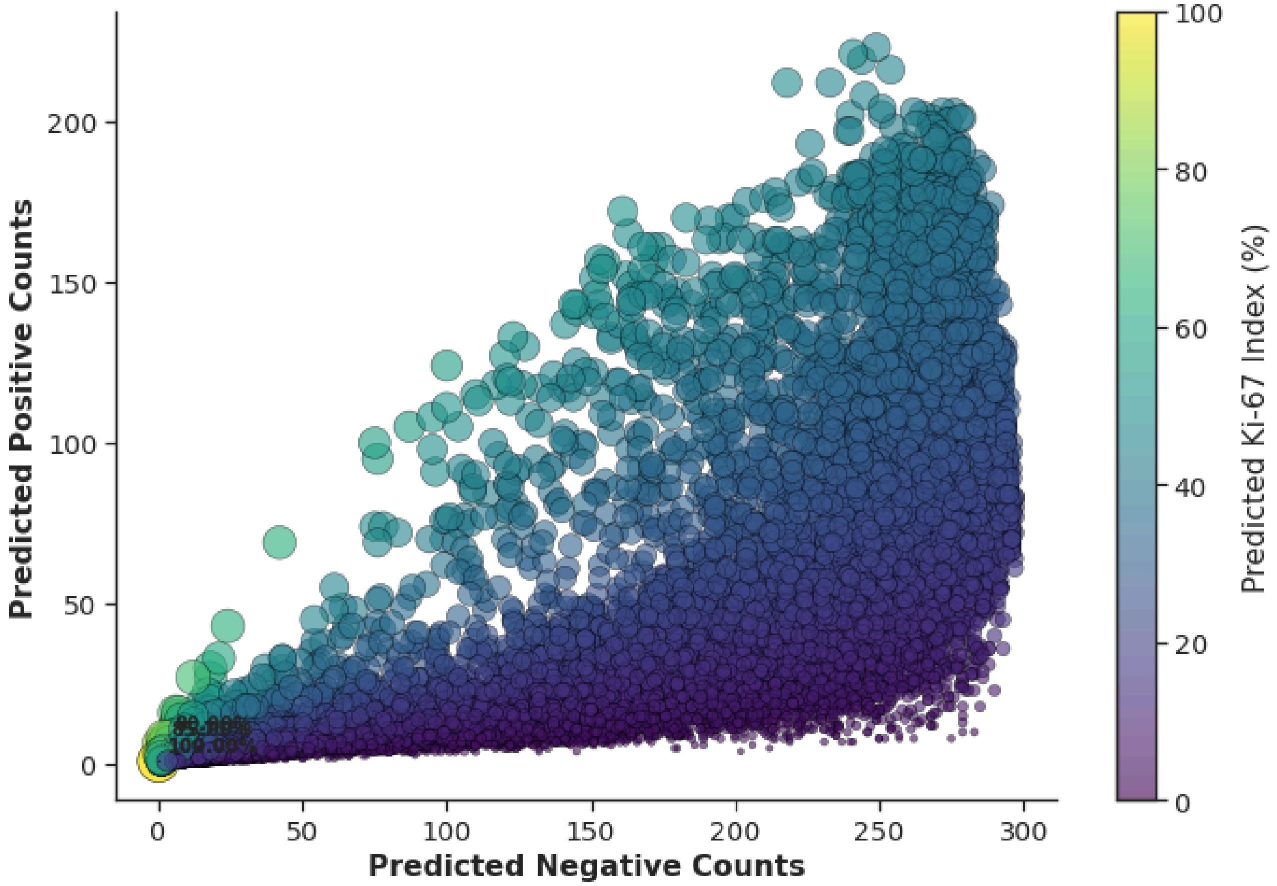


**Supplementary Figure S8.** *The scatter plot shows the relationship between predicted Ki-67-positive and Ki-67-negative cell counts. Point size reflects the total predicted cell count, while colour suggests the predicted Ki-67 index (%) from the IHC4BC dataset.*

**Supplementary Table S3.** *Performance of the cross-modality model on the testing set.*

| Class | MAE (95% CI) | RMSE (95% CI) | $R^2$ (95% CI) |
|---|---|---|---|
| **Positive Cells** | 4.68 (4.46, 4.89) | 8.49 ( 8.09, 8.89) | 0.95( 0.95, 0.96) |
| **Negative Cells** | 11. 08 ( 10.70, 11.47) | 17.04( 16.38, 17.65) | 0.97(0.97, 0.98) |
| **Ki-67 Index** | 3.25 (3.14, 3.38) | 5.06 ( 4.80, 5.33) | 0.80 (0.78, 0.82) |

**Abbreviations:** MAE: Mean Absolute Error; RMSE: Root Mean Squared Error; $R^2$: Coefficient of Determination; CI: Confidence Intervals.

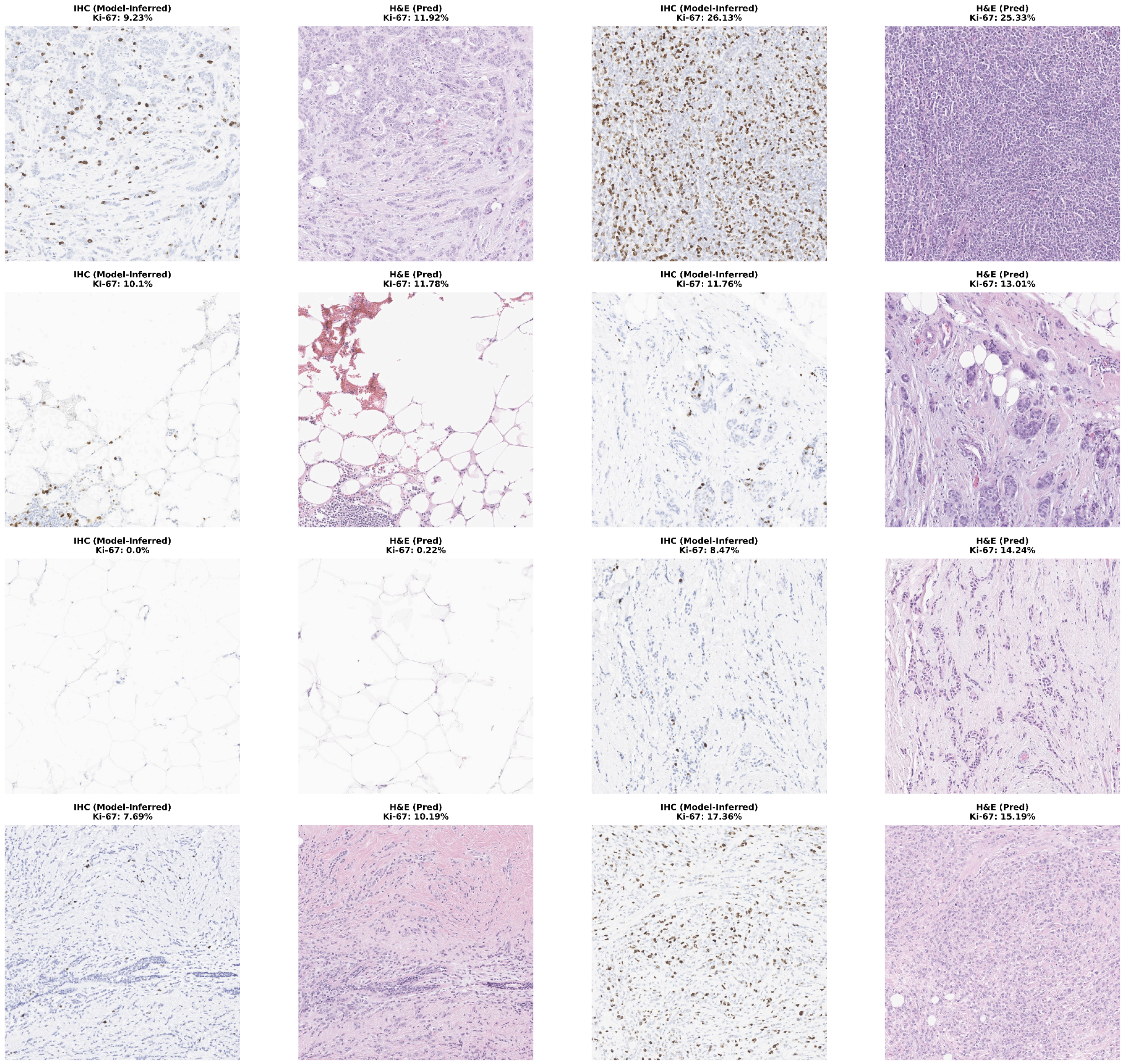


**Supplementary Figure S9.** *Visualisation of the Ki-67 index from paired IHC and H&E-stained patches is presented as follows: (IHC → H&E) | (IHC → H&E). The IHC patches show the Ki-67 index as inferred from the IHC regression model. In contrast, the corresponding H&E patches display the Ki-67 index, which is quantified using the cross-modality H&E regression model.*

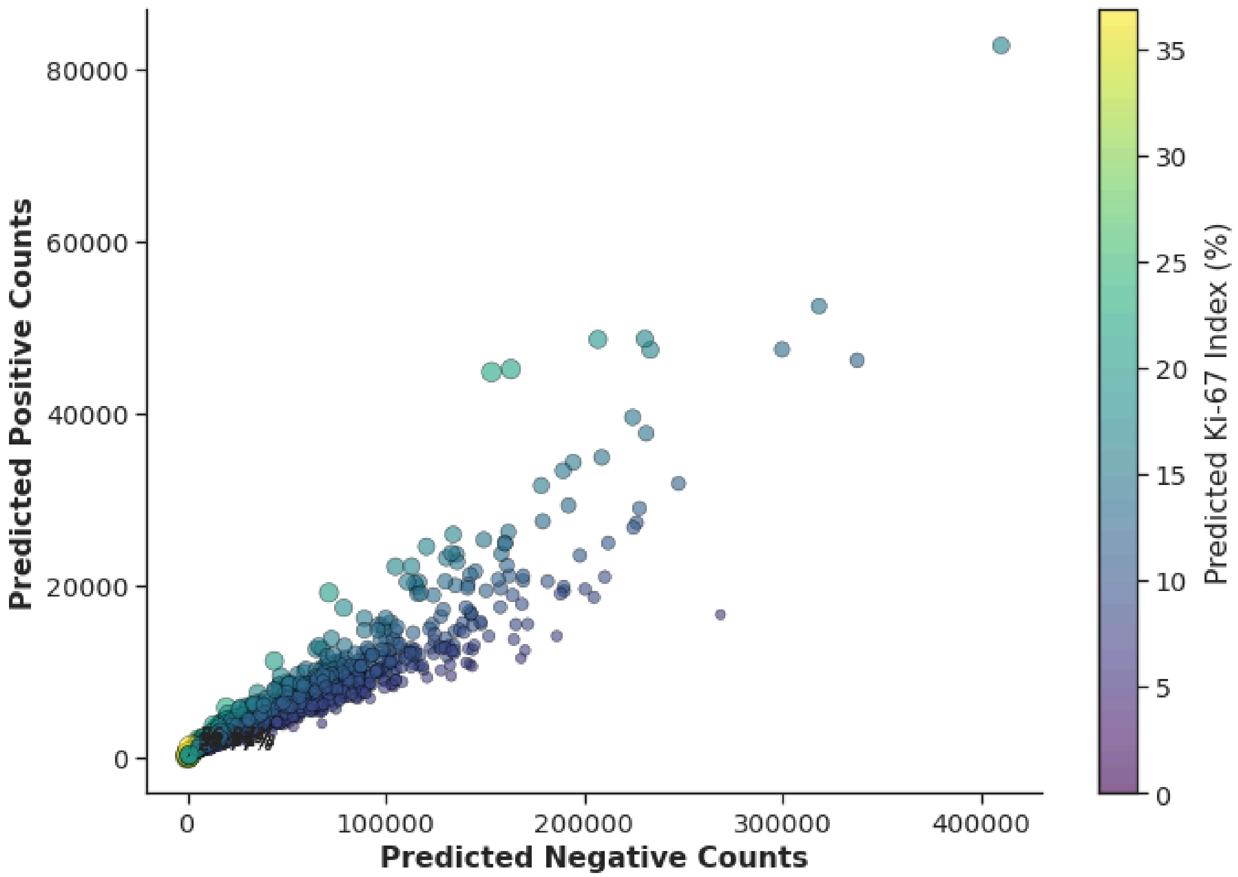


**Supplementary Figure S10.** *Predicted counts of Ki-67-positive and negative cells using the regression model across 846 IHC-stained whole slide images (WSIs) from the ACROBAT dataset. The size and colour of the bubbles represent the slide-level Ki-67 index (%).*

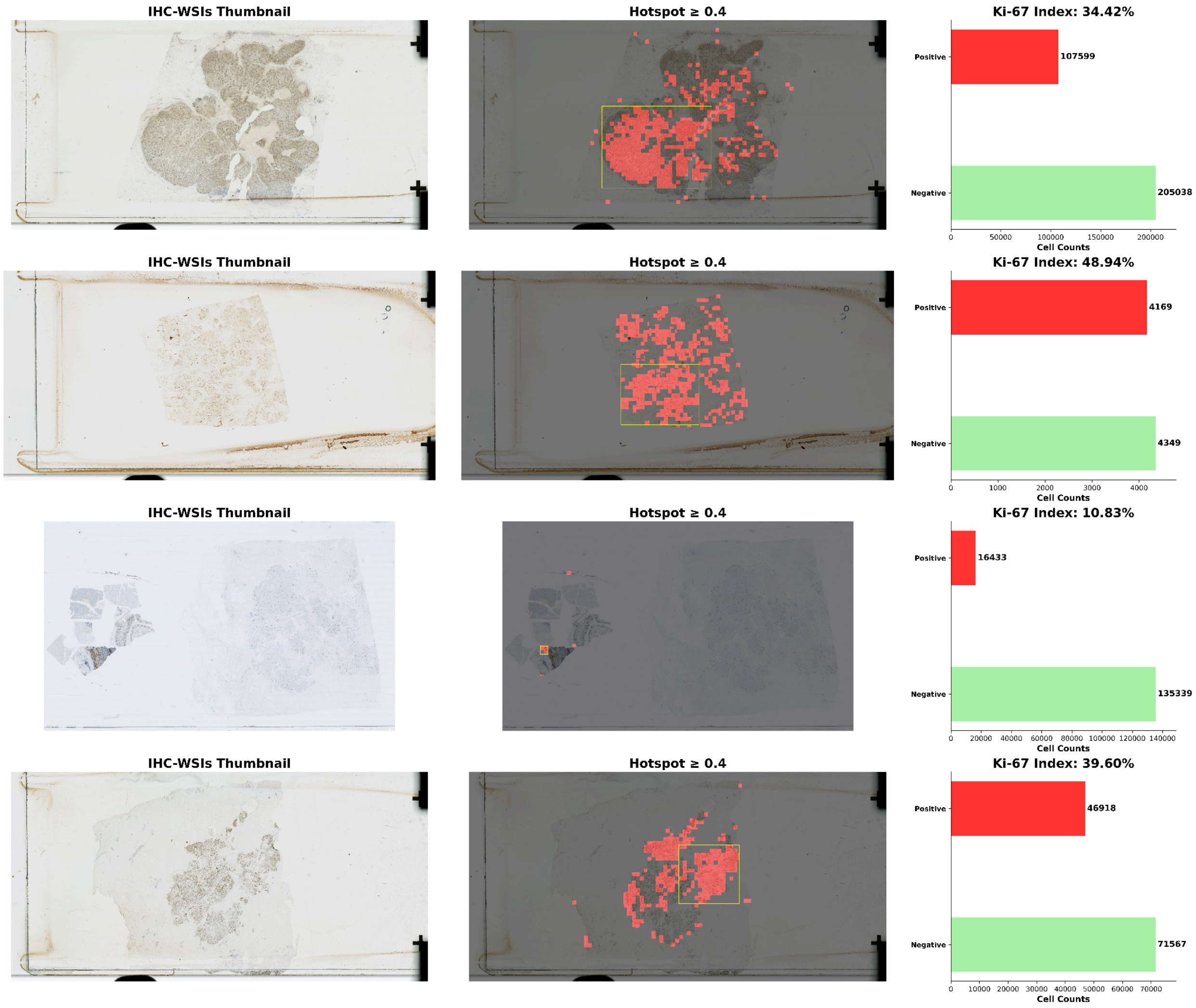


**Supplementary Figure S11.** *Quantification of Ki-67 index at whole slide level and visualisation of hotspots using the ACROBAT dataset. Four slide images with differing predicted Ki-67 indices were selected. For each slide, the left side displays the original thumbnail of IHC-stained WSIs, the middle panel overlays predicted Ki-67 hotspots in red, with the largest hotspot region outlined (yellow), and the right panel presents bar plots of predicted Ki-67-positive (red) and Ki-67-negative (green) cell counts.*

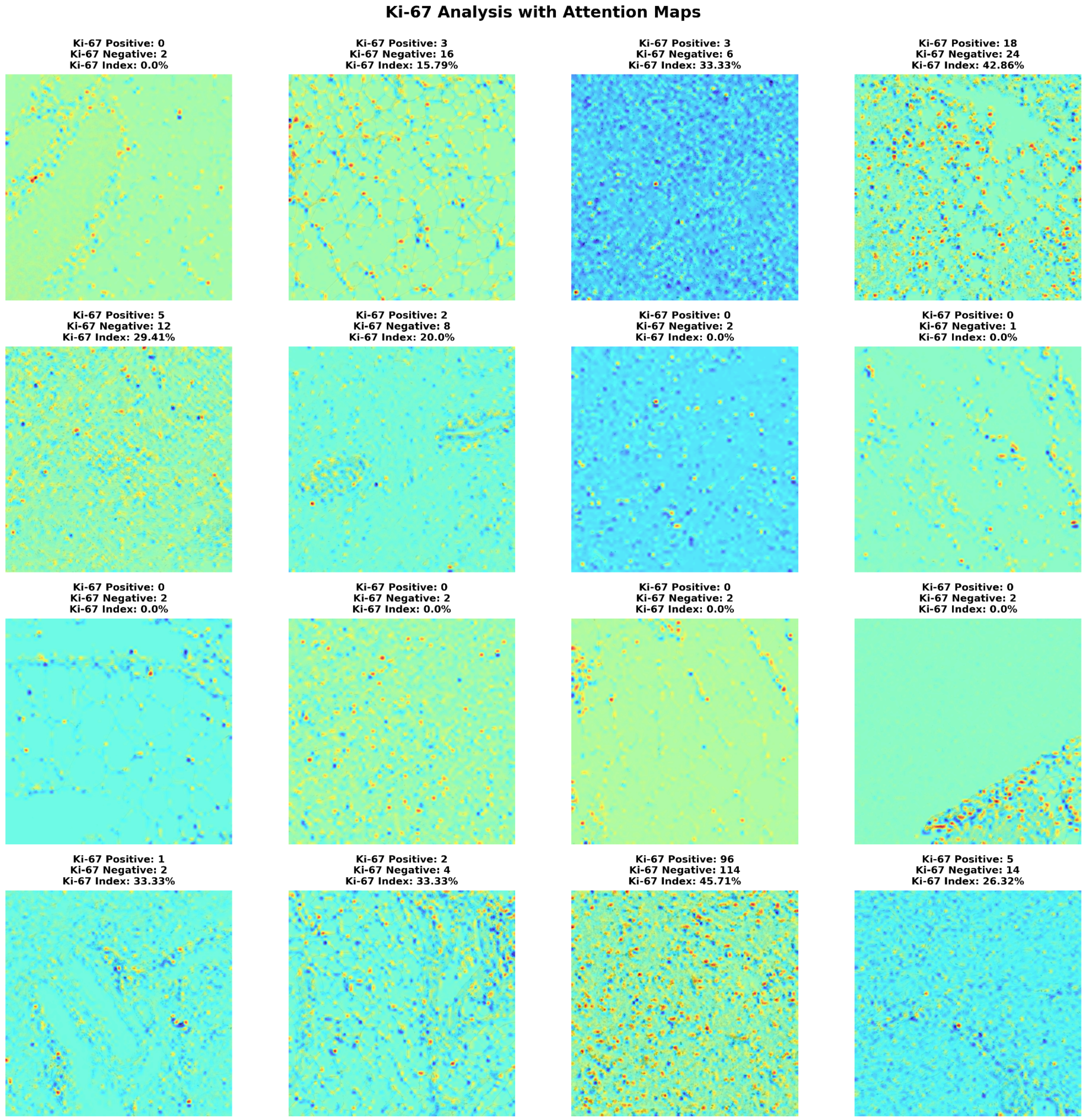


**Supplementary Figure S12.** *Visualisation of randomly predicted Ki-67 cell counts from the IHC4BC dataset.*

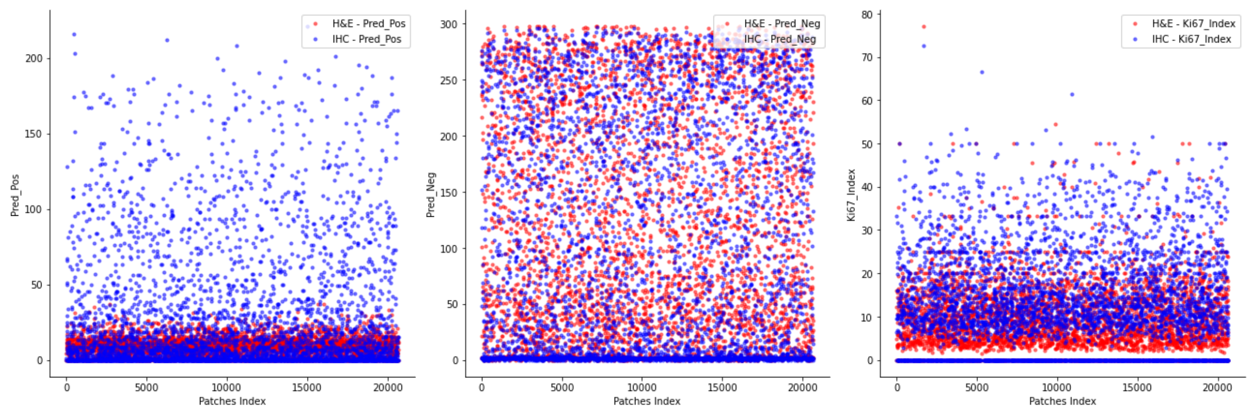


**Supplementary Figure S13.** Predicted Ki-67 counts on H&E and IHC-stained patches across all the testing sets using our regression model. The left and middle panels display predicted counts of positive and negative cells, respectively, while the right panel displays Ki-67 index values. The middle panel illustrates a notable overlap in negative cell counts between IHC (blue) and H&E (red) stained patches, indicating a high level of consistency. The right panel shows that Ki-67 index values from H&E-stained patches are generally lower than those from IHC-stained patches, highlighting a consistent underestimation of Ki-67 expression from H&E-stained patches.
